# Machine learning-based prediction of motor status in glioma patients using diffusion MRI metrics along the corticospinal tract

**DOI:** 10.1101/2021.08.24.21262484

**Authors:** Boshra Shams, Ziqian Wang, Timo Roine, Baran Aydogan, Peter Vajkoczy, Christoph Lippert, Thomas Picht, Lucius S. Fekonja

## Abstract

Along tract statistics enables white matter characterization using various diffusion MRI (dMRI) metrics. Here, we applied a machine learning (ML) method to assess the clinical utility of dMRI metrics along corticospinal tracts (CST), investigating whether motor glioma patients can be classified with respect to their motor status. The ML-based analysis included developing models based on support vector machine (SVM) using histogram-based measures of dMRI-based tract profiles (e.g., mean, standard deviation, kurtosis and skewness), following a recursive feature elimination (RFE) method based on SVM (SVM-RFE). Our model achieved high performance (74% sensitivity, 75% specificity, 74% overall accuracy and 77% AUC). Incorporating the patients’ demographics and clinical features such as age, tumor WHO grade, tumor location, gender and resting motor threshold (RMT) into our designed models demonstrated that these features were not as effective as microstructural measures. The results revealed that ADC, FA and RD contributed more than other features to the model.

## Introduction

Gliomas are known as the most frequent and malignant human brain tumors, characterized by poor prognosis and high morbidity ^1^. Gliomas infiltrating the motor system potentially cause various degrees of damage to the white matter (WM) architecture and might lead to substantial motor function impairments. Diffusion-weighted imaging (dMRI) ^2, 3^ has shown promising potentials by enabling non-invasive delineation of the WM pathways known as tractography ^4–8^. Tractography has been frequently used for preoperative planning or analyzing tumor-induced structural impact, for example to investigate tumor infiltration and its impact on surrounding tissues ^9, 10^. Here, we investigate whether dMRI-based metrics can be used as predictive features to detect motor function impairments.

Recent studies demonstrated clinical correlations of segmental diffusion tensor imaging (DTI) derived metrics, such as apparent diffusion coefficient (ADC; a measure of the overall diffusivity in a single voxel), axial diffusivity (AD; the diffusion rate along the main axis of diffusion), fractional anisotropy (FA; the directional preference of diffusion), or radial diffusivity (RD; rate of diffusion in the transverse direction) ^11–13^. Furthermore, we investigated how a more complex dMRI-based metric, namely the constrained spherical deconvolution (CSD)-related fiber density (FD) offers an even more detailed WM characterization ^14^. DTI and CSD-related quantification of dMRI-based measures along tractograms ^15^ have gained great interest since these methods reveal insights into WM development, function and disease ^16–19^. Multiple fiber populations are found in up to 90% of the WM voxels and 30% - 40% of these WM voxels contain more than three fiber populations ^20–23^. Moreover, non-WM contamination is found in more than a third of the WM voxels ^24^ and multi-tissue CSD methods ^25–28^ have been used to account for it. As a result, CSD-based metrics in addition to DTI metrics (such as AD, ADC, FA, or RD) are critical. By estimating fiber orientation distributions (FODs) in each voxel based on the expected signal from a single collinearly oriented fiber population, CSD can discriminate complex fiber populations ^29^. Probabilistic tractography algorithms, such as the iFOD2, have been proposed to overcome the limitations of tensor-based tractography methods by using the rich information in FODs ^30^. A complete picture of the underlying white matter architecture is critical for risk assessment and neurosurgical planning and as well for prediction models^31^. To that end, modern CSD-based FD and fixel-based analysis (FBA) approaches, in addition to traditional DTI methods, provide promising opportunities because they are related to the intra-axonal restricted compartment that is limited to a given fiber orientation within a voxel ^32, 33^. Recently, we used FD for fiber orientation-specific study of microstructural properties along the tract in relation to infiltrating tumors ^14^, which was previously focused on group-based analyses ^32^. Yet, research lacks the individual and tract-specific characterization of white matter microstructure investigating the association between tumor impact on structural connectivity and clinical outcomes.

Here, we employed machine learning (ML) methods using along CST-related dMRI metrics (e.g., AD, ADC, FA, FD and RD) to predict motor deficits in patients with motor-related glioma, focusing on individual diagnosis rather than groupwise comparisons. We used ML methods based on support vector machines (SVM) which is a powerful method, easy to interpret and well suitable method to handle large dimensional datasets ^34–36^. We designed our predictive models applying SVM with an embedded feature selection methods ^37, 38^ using histogram-based features of dMRI-based tract profiles. In addition, an SVM model was developed with principle component analysis (PCA) method ^39, 40^ to process all segmental information of dMRI-based tract profiles in a low dimensional feature space. Furthermore, patients’ demographics and clinical variables including the resting motor threshold (RMT), a transcranial magnetic stimulation (TMS)-derived neurophysiological marker, were incorporated into our designed models, sought to investigate the impact of all types of features, e.g., demographics, clinical and microstructural features, on the performance of the developed models.

## Results

MRC grading, TMS mapping to acquire the RMT, the computation of CST and the subsequent extraction of tractogram-related AD, ADC, FA, FD and RD measures were feasible in each patient. Visual inspection of violin- and line plots showed differences between patients with (class 1) and without (class 0) motor deficits in ipsilesional tractograms for AD, ADC, FA, FD and RD (*Fig. 1 & 2*). 45 (37.9%) of the recruited 116 patients presented with preoperative motor deficits (MRC<5), cf. *Table 1*. There were no significant differences in gender (χ^2^[1, N = 116] = 0.51, p = 6.3e − 1) or hemispheric pathology position (χ^2^[1, N = 116] = 0.08, p = 7.7e − 1) in relation to motor deficits. Patients with motor deficits (class 1) were older (58.64 ± 15.45) than patients without motor deficits (class 0) (50.25 ± 15.85), with a highly significant difference between them (t[114] = 2.83, p = 5e − 3), although a medium effect was found (g = 5.3e − 1 95% CI = [.155 − .914]). There were no significant differences in tumor locations and RMT ratio in both ipsi- and contralesional hemispheres in relation to motor deficits, cf. *Table 1*. We also found significant difference between the glioma WHO grade III and IV between patients with and without motor deficits ( χ^2^[1, N = 116] = 0.86, p = 3e − 2).

**Figure 1:**
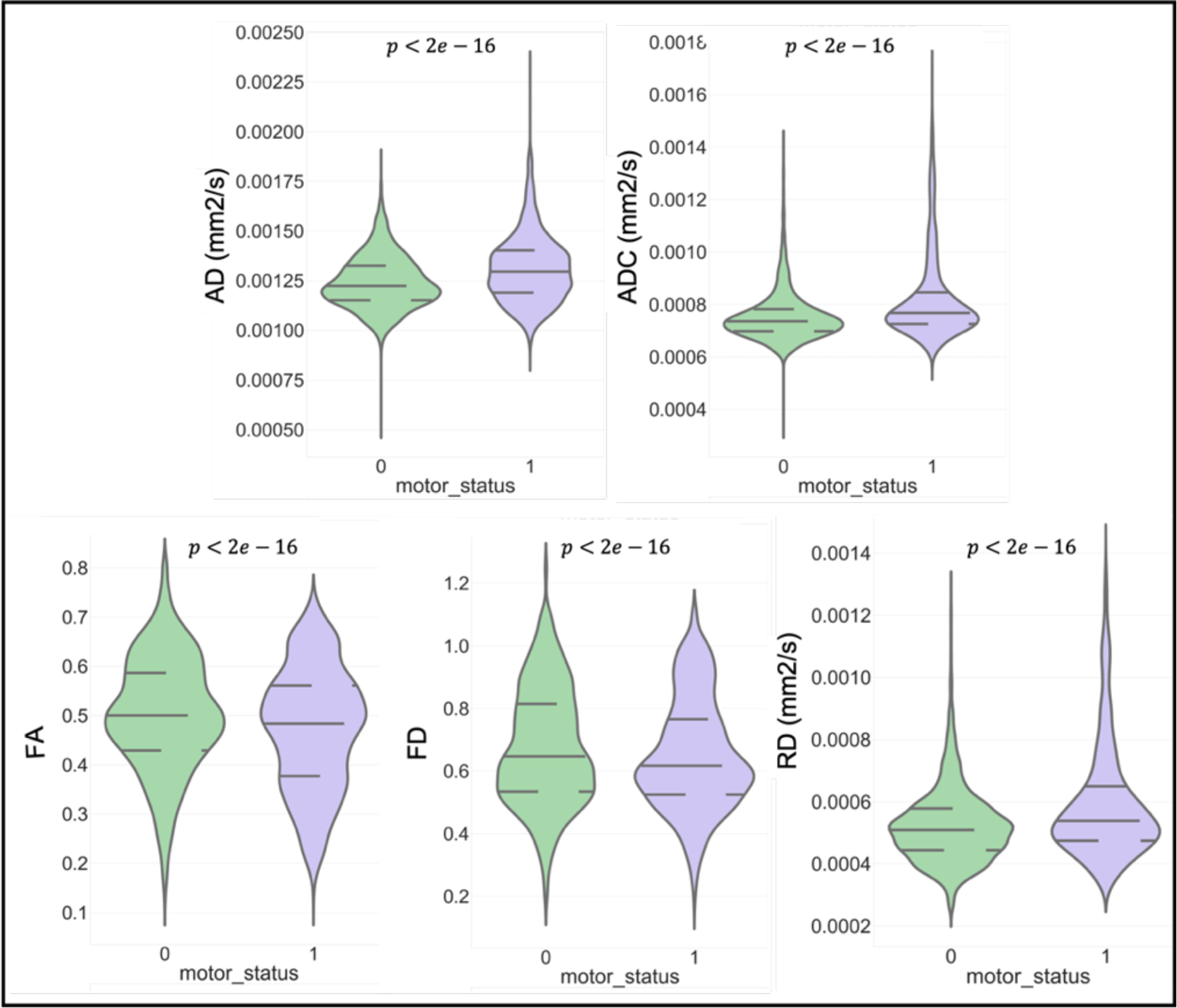
Violin plots illustrating frequency distribution of AD, ADC, FA, FD and RD metrics over the ipsilesional CST (class 0, without motor deficit, green, MRC = 5; class 1, with motor deficit, violet, MRC<5); horizontal lines indicate quartile positions.

**Table 1.** Demographic and neuropathological overview of the patient cohort

|  | Patients without motor deficits<br>Class 0 | Patients with motor deficits<br>Class 1 | *p-value* |
| --- | --- | --- | --- |
| **Demographics** |  |  |  |
| Cohort size | 71 (62%) | 45 (37%) | – |
| Age | 50.25±15.85 | 58.64±15.45 | 0.005 |
| Female | 25 (35%) | 18 (41%) | 0.63 |
| Male | 47 (65%) | 26 (60%) | 0.63 |
| **Ipsilesional hemisphere** |  |  |  |
| Left | 30 (42%) | 17 (39%) | 0.77 |
| Right | 41 (57%) | 28 (64%) | 0.77 |
| **Tumor location** |  |  |  |
| Frontal | 35 (49%) | 25 (55%) | 0.55 |
| Parietal | 19 (27%) | 12 (27%) | 0.91 |
| Insular | 10 (14%) | 5 (11%) | 0.77 |
| Temporal | 7 (10%) | 3 (7%) | 0.73 |
| **Glioma WHO grades** |  |  |  |
| II | 13 (18%) | 3 (7%) | 0.8<sup>a</sup>, 0.05<sup>b</sup> |
| III | 18 (25%) | 5 (11%) | 0.8<sup>a</sup>, 0.03<sup>c</sup> |

| IV | 40 (55%) | 37 (84%) | 0.05<sup>b</sup>, 0.03<sup>c</sup> |
| --- | --- | --- | --- |
| **RMT (V/m)** |  |  |  |
| Ipsilesional hemisphere | 33.72±7.2 | 34.64±9.15 | 0.57 |
| Contralesional hemisphere | 34.3±6.05 | 35.62±8.64 | 0.37 |

\* Resting Motor Threshold (RMT), TMS-derived neurophysiological marker, indicating cortical excitability; <sup>a</sup> Tumor type II versus III; <sup>b</sup> Tumor type II versus IV; <sup>c</sup> Tumor type III versus IV.

Moreover, considering CCA, we found a strong positive correlation between all dMRI-based extracted features and age, which was statistically significant (r_s_(114) = 0.7, p = 3.3e − 18). The correlation coefficient is provided in supplementary table 1. This correlation was stronger in the group with motor deficits (class 1) (r_s_(45) = 0.93, p = 6.12e − 21). Considering each metrics separately, the correlation was weaker (**AD:** r_s_(45) = −0.59, p = 1.64e − 5; **ADC:** r_s_(45) = 0.64, p = 2.32e − 6; **FA:** r_s_(45) = −0.42, p = 3.2e − 4; **FD:** r_s_(45) = −0.24, p = 0.1; **RD:** r_s_(45) = 0.51, p = 3.54e − 4). In the group without motor deficits (class 0), the dMRI-based extracted measures were negatively correlated to age (r_s_(71) = −0.89, p = 1.0e − 25). Considering each metrics separately, the correlation was weaker (**AD:** r_s_(71) = 0.33, p = 4.0e − 4; **ADC:** r_s_(71) = 0.53, p = 2.07e − 6; **FA:** r_s_(71) = −0.49, p = 1.64e − 5; **FD:** r_s_(71) = −0.55, p = 4.87e − 7; **RD:** r_s_(71) = 0.58, p = 1.2e − 7).

### Group-wise statistical analysis

A comprehensive group-wise analysis was performed over the entire CST and segment-wise along the 100 segments to compare the differences of dMRI metrics between the two patients’ groups with (class 1) and without (class 0) motor deficits. The violin plots in *Fig. 1* shows significant differences in dMRI-based measures in the ipsilesional CST between the two patients’ groups.

*Fig. 2* shows a segment-wise comparison of dMRI metrics between the two groups of patients (class 0; class 1) in ipsilesional CST. We found significant segment-wise differences, surviving false discovery rate (FDR) correction, between the two groups in the ipsilesional CST profiles in relation to ADC, AD, FA and RD metrics. However, no significant segment-wise differences were found with respect to FD (cf. supplementary table 2). These differences were larger in ADC and RD, especially in the tracts’ middle and peritumoral areas. The ipsilesional tract profiles led to larger significant differences compared to the differences between ipsi- and contralesional CSTs in group analyses (cf. *supplementary table 3*). In the latter case, only 6 nodes (93-98th) in the ADC, 9 nodes (88-96) in FA and 11 nodes (88-98) in RD metrics of the ipsilesional tracts showed significant differences between the two groups (class 0; class 1) mainly at the superior portion of the CST.

**Figure 2:**
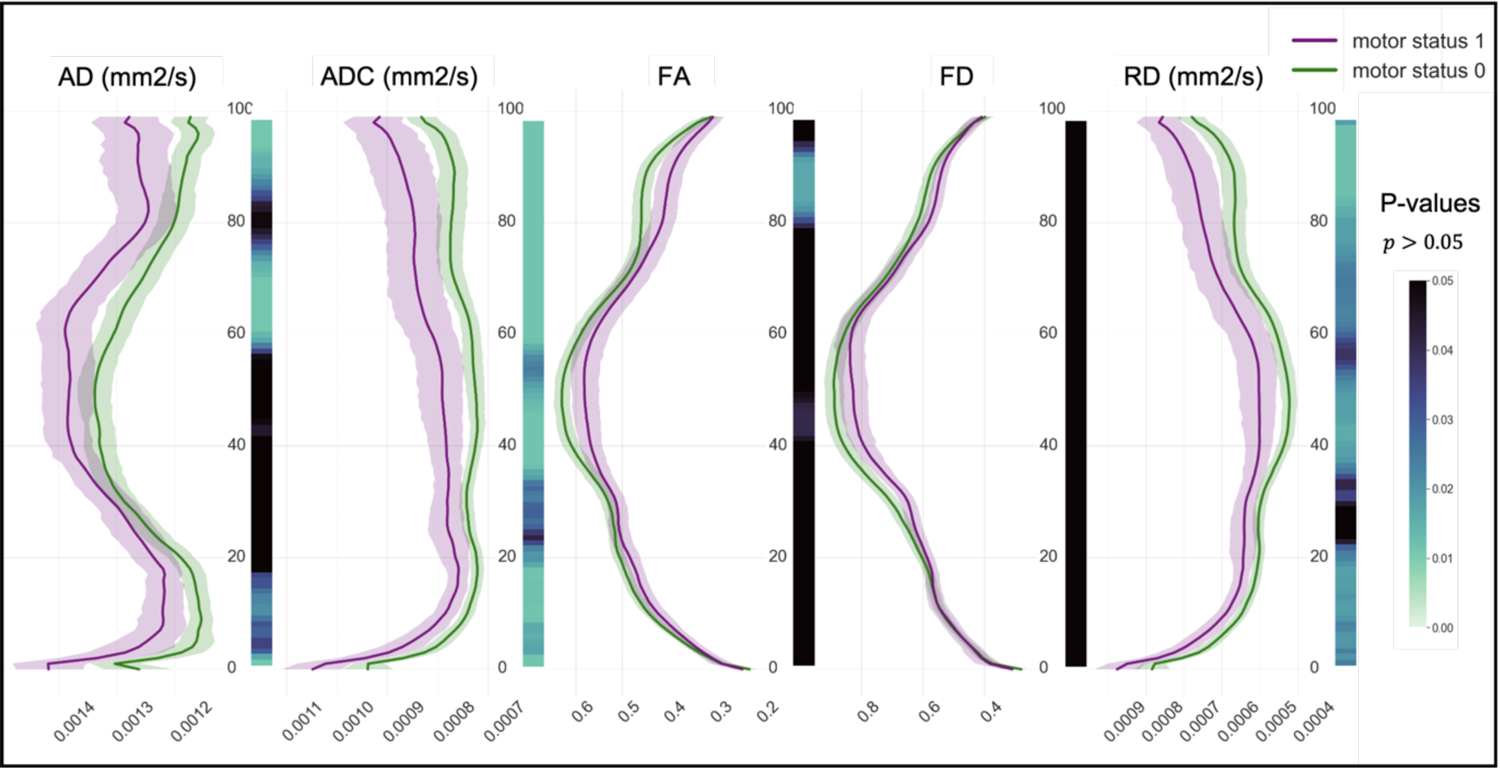
Line plots illustrating AD, ADC, FA, FD and RD metrics along the ipsilesional tractogram (segment 0 = medulla oblongata; segment 100 = M1), for both motor patients’ groups (class 0, green, MRC = 5; class 1, violet, MRC<5). The lines indicate median values with their 95% confidence interval. The heatmaps demonstrate related FDR-BH corrected p-values, derived by t-test.

**Table 2.** SVM_1-4 model performances using Mdn vs. KNN interpolation methods and Mdn vs. and Mahalanobis-based weighted mean tract profile.

|  | Accuracy<br>(%) | Sensitivity<br>(%) | Specificity<br>(%) | AUC<br>(%) |
| --- | --- | --- | --- | --- |
| **Mdn-based Profile** |  |  |  |  |
| Mdn interpolation (SVM_1) | 73 | 73 | 73 | 76 |
| KNN interpolation (k = 10) (SVM_2) | **74** | **74** | **75** | **77** |
| **Weighted M profile** |  |  |  |  |
| Mdn interpolation (SVM_3) | 68 | 66 | 70 | 70 |
| KNN interpolation (k = 10) (SVM_4) | 69 | 66 | 70 | 70 |

**Table 3.** SVM2 selected features with their respective learned weights using SVM-RFE.

| Features | Weights |
| --- | --- |
| FA_KU | 1.29 |
| FA_SK | 1.17 |
| RD_KU | 1.07 |
| ADC_STD | 1.01 |
| ADC_M | 0.61 |

Moreover, the significant segments in contralesional tract profiles were only seen in few segments in AD and ADC tract profiles (cf. *supplementary table 4*).

Additionally, we used M, STD, KU and SK as histogram-based measures of tract profiles and performed group-wise analyses on each measure for ipsi- and contralesional tract profile (cf. *supplementary table 5a* & *5b*). As shown in *Fig. 3 (A, B, C, E)*, the M measure over the ipsilesional CST profiles of AD, ADC, FA and RD were significantly different between the two patients’ groups with and without motor deficits (**AD:** U = 1000, p = 3.6e − 4, r = 0.37; **ADC:** U = 974, p = 2.07e − 4, r = 0.4; **FA:** U = 1214, p = 1.5e − 2, r = 0.24; **RD:** U = 1003, p = 3.8e − 4, r = 0.42) as well as the KU measure of ipsilesional CST profile of FA value (U = 1034, p = 7.1e − 4, r = 0.35). Further in *Fig. 3 (B, E*), STD of ADC and RD profiles showed highly significant increase in patient group with motor deficit (**ADC:** U = 1032, p = 6.8e − 4, r = 0.35; **RD:** U = 1065, p = 1.2e − 3, r = 0.33).

**Figure 3:**
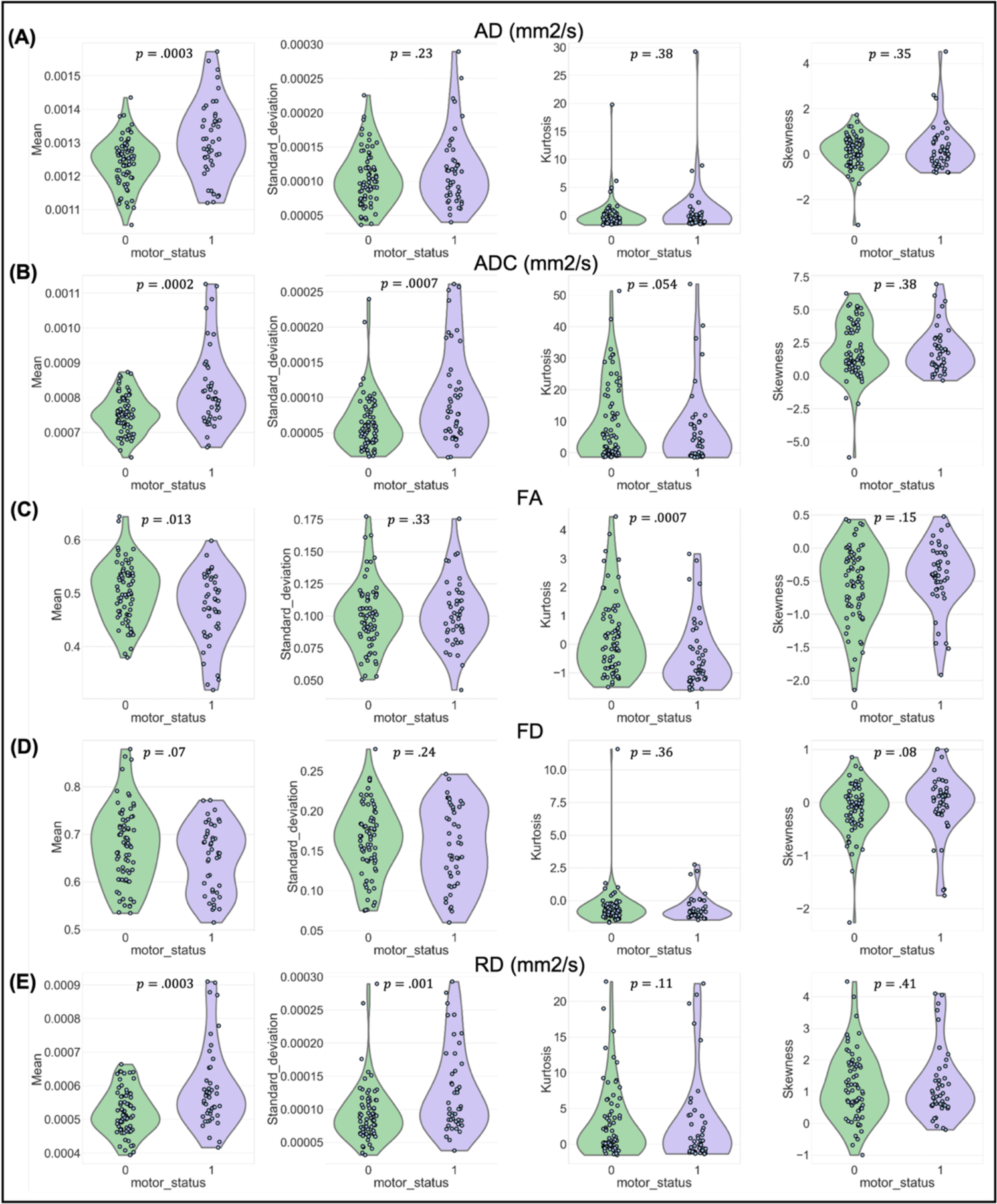
Violin plots illustrating different measures of CST profiles for AD (mm2/s), ADC (mm2/s), FA, FD and RD (mm2/s) metrics. The analyses were done in ipsilesional tract (affected hemisphere) profiles between two patient groups, with and without motor deficits (class 1, green, MRC <5; class 0, violet, MRC=5).

Ipsilesional tract profile measures led to larger differences compare to the contralesional tract profile measures. Nine ipsilesional tract profile measures (extracted features) were significantly different between the two patients’ groups (cf. *supplementary table 5a*), see *Fig. 3*, while only three measures of contralesional tract profiles were significantly different (cf. *supplementary table 5b*).

### SVM classification

SVM_clinical, using all patients’ demographic and clinical variables as input features, received low performance score (58% accuracy, 82% sensitivity, 43% specificity and 62% AUC). Among all SVM_AD-RD models, when using microstructural measures in relation to each metric separately, SVM_FA reached to the highest accuracy (67%), sensitivity (60%) and AUC (68%) and SVM_ADC and SVM_RD reached to the highest specificity (80%). The AUC in SVM_ADC and SVM_AD reached to nearly the same score, 67%, *Fig. 4*.

**Figure 4.**
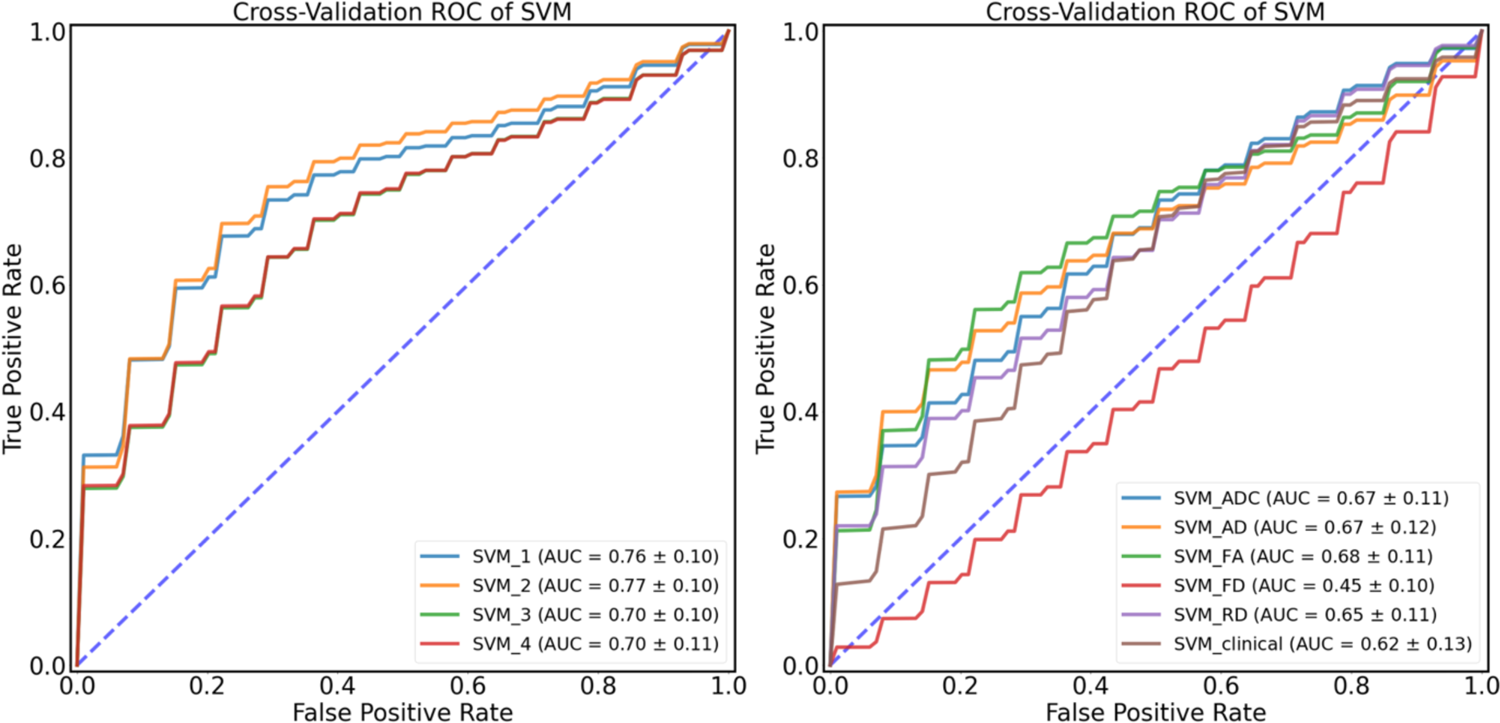
Receiver operating characteristic (ROC) curves of the discriminative performance of the SVM_1-4 and SVM_ADC-RD models show the true positive rate against the false positive rate for different threshold; The blue lines indicate classifiers with a random performance level.

Considering SVM_1-4 models, using all measures of dMRI-based tract profiles, the best classifier performance was achieved with SVM_2, which reached to 74% accuracy, 74% sensitivity, 75% specificity and 77% AUC (*Table 2)*. With KNN interpolation method, the number of nearest neighbor when K=10 yielded the best model performance.

The 5 most effective features that were selected by SVM-RFE are FA_KU, FA_SK, RD_KU, ADC_STD and ADC_M; effect sizes and FDR-corrected *p*-values were also calculated (**FA_KU:** U = 1034, p = 1.1e − 3, r = 0.35; **FA_SK:** U = 1413, p = 1.5e − 1, r = 0.11; **RD_KU:** U = 1377, p = 1.3e − 1, r = 0.13; **ADC_STD:** U = 1032, p = 1.1e − 3, r = 0.35; **ADC_M:** U = 974, p = 1.0e − 3, r = 0.4). *Table 3* shows these selected features with their respective learned weights using SVM_2.

In the last step, patients’ demographics and clinical features were integrated into our models (SVM_1-4). The feature selection method did not select them in any of the trained models (SVM_1-4) and the models’ performances remained unchanged. The receiver operating characteristic curves (ROC) for SVM_1-4, SVM_AD-RD and SVM_clinical are presented in *Fig. 4*.

Moreover, we developed a model using all segment-wise information along ipsi- and contralesional tract profiles for all dMRI metrics (ADC, RD, AD, FA and FD) with PCA(SVM_5). This model reached to 63% accuracy, 64% sensitivity, 63% specificity and 70% AUC.

## Discussion

We investigated how glioma-induced microstructural alterations to WM associated with functional motor deficits by mapping dMRI metrics along the CST. In segment-wise group comparison, we found significant differences between the ipsilesional tract profiles in the two patients’ group (class 0; class 1) in relation to all dMRI metrics except for FD. The ipsilateral differences were mainly seen at the level of the glioma (superior portion of the tract), which showed the direct influence of the tumor area on motor function. However, tumor impact on tract metrics could also be detected in areas relatively distant from the tumor and peritumoral edema especially in ADC and RD, demonstrating the spreading of the local tumor effect with respect to the motor status.

FA and ADC have been widely utilized to evaluate the WM health to large extent. FA has been shown to be highly sensitive for detecting changes in WM water diffusion in case of neuropathology, but rather unspecific. An increase in ADC has been observed in pathologies accompanied by for example edema or necrosis ^30, 41^. However, these two measures are less sensitive to distinguish specific types of neural impairments (e.g., demyelination, axonal injury, inflammation). Since the majority of our patient’s cohort encompassed patients with glioblastoma tumors (77 patients) with the hallmark of diffuse infiltration into the normal brain, we could see the greatest elevation of ADC values along CST compared to the other metrics. These results confirmed that the myelinated ipsilesional CST fibers were affected.

Furthermore, AD and RD are direct measures of diffusion in perpendicular and in parallel directions to the tract, respectively. The incorporation of these metrics has been shown to lead to better differentiations between axonal injury or degeneration (AD) and pathological demyelination (RD) ^30, 41–44^. Demyelination could be more a chronic/slow process, while axonal degeneration could be a more acute and potentially clinically more detrimental process. Here, we did not see a strong improvement in our SVM models performance on basis of AD. However, the Kurtosis of tract profile on the basis of RD was highly effective (see below).

Besides, we even found differences in segments/regions along contralesional tract profiles with respect to ADC and AD, supporting the assumption that contralesional CST may play a role in postoperative motor outcome and in recovery of motor function. These results may have implications for compensatory strategy of the brain.

### SVM analysis Microstructural measures

Considering dMRI-based features within the ipsi- and contralesional CST, we successfully developed several SVM-based models to classify patients with respect to their motor deficits (class 0; class 1). The best model performance achieved an AUC of 76.6% (SVM_2). The most effective features were FA_KU, ADC_M, RD_KU, FA_SK and ADC_STD. Accordingly, ADC and FA were identified as the most relevant metrics which better accounted for detection of motor function impairment ^45^. These findings are also in line with our previous studies ^13, 14^. In ^14^, we found FA and ADC metrics to be the most relevant metrics for detecting CST impairments, and in ^13^, we found that peritumoral ADC and FA were strongly associated with postoperative motor deficits (motor outcome). The univariate analysis confirmed our SVM model results to some extent, showing the predictive power of each tract profile’s measure (input feature) separately. All models which were trained on each AD, ADC, FA and AD metrics separately, reached a relatively high specificity though they dealt with low sensitivity. Among them, SVM_FA provides a higher sensitivity but lower specificity and SVM_ADC and SVM_RD provided higher specificity. Combining these metrics (SVM_2) led to both high sensitivity and specificity. FA_SK and RD_KU were not significantly different between the two patient groups although they were selected as two of the most effective features. Features identified as significantly relevant or/and predictively relevant can agree or diverge, and numerous studies have been conducted demonstrating the differences between highly predictive and highly significant variable sets ^46, 47^. Sometimes a strong predictivity fails to be significant, as it only provides supplementary information and could increase the predictive power of ML models just in combination with other features. The SVM_2 model reached the best performance among all trained models which showed that the Mdn profile corresponded to a better prediction accuracy in comparison to the Mahalanobis-based weighted M tract profiles – a method described in ^17, 48^, that implies the robustness of the Mdn method in the specific case, since outlier segments with extreme values do not bias the Mdn. The superiority of the Mdn profile has been previously shown in ^49^, where microstructural models were used to understand the role of WM in relation to cognitive development. Our SVM_1-4 models, which were based on the extracted features, were more efficient and robust compared to SVM_5 since these models were less complex in their design. The high dimensional dMRI data (100 values per tract profile) and low available number of patient data resulted in lower performance in SVM_5. The variety in glioma location in relation to the CST as well as the low sample size restricted our analysis when SVM_5 tried to capture all patterns of microstructural variations. Therefore, we were not able to detect all segment-wise variations as efficiently as possible in SVM_5. In the SVM_1-4 models, we were able to summarize the segment-wise information of dMRI-based CST profiles as different statistical measures to detect informative glioma-induced microstructural alterations to WM to predict functional motor deficits.

### Demographic and clinical features

The SVM model with only demographics and clinical variables (SVM_clinical) such as age, gender, glioma location, glioma WHO grade and RMT ratio, showed a poor performance (56% accuracy and 62% AUC) while the models with microstructural measures as input, e.g., SVM_2 (74% accuracy and 77% AUC), reached to relatively high performance. We further assessed the performance of the SVM_1-5 models integrating patients’ demographics and clinical features and saw that none of them was affected the SVM models’ performances. This could indicate a lower effectiveness of these features compared to tract profile-based characteristics (microstructural measures). In addition, the dMRI-based measures could be associated with different variables. Since patient’s age was significantly different between the two patients’ groups, it was expected to improve our predictive models’ performances in combination with the microstructural measures. However, integration of patients’ age did not improve the models’ performances. The performed CCA showed a strong and significant correlation between dMRI extracted features and age. Interestingly, we found strong and significant correlation in the reverse direction for both motor groups (class 0; class 1). This confirms previous findings ^50–52^ in which WM changes in relation to age and its variation as a function of age were investigated. In a recent study, age has been accurately predicted by FA and ADC metrics^48^. These results justify as well that if taking into account the microstructural measures, age is of critical importance in distinguishing between the two motor groups.

According to CCA analysis and our SVM results, we could conclude that the information provided with age was apparently sufficiently covered by dMRI-based extracted features and thus no additional information was found considering patients’ age.

### Translational aspect

The body of evidence that preservation of the white matter connectivity is key to preserving function is steadily growing. Therefore, the presurgical assessment not only of the spatial relation of the tumor and the tracts, but also a detailed analysis of the impact the tumor already exerts on the white matter is of great importance. ML, as demonstrated in this study, shows a promising potential to address the microstructural effects of brain tumors on the WM which is not accessible with traditional statistical methods, since it allows for discovering patterns in dMRI data and well approximating complex relationships. Future studies need to further correlate ML finding with functional outcomes to establish new biomarkers for WM resilience to surgical manipulation with the promise to become a powerful prognostic tool in future neurosurgery.

### Limitations

Tractography suffers from a wide range of limitations that make its routine use problematic ^14, 53^. Tractograms contain both false positive ^54^ and false negative ^55^ streamlines. In addition, tractography cannot distinguish between afferent and efferent connections, and streamlines may terminate improperly ^56^, especially in case of tumors and edema. Furthermore, our results are atlas and tractography algorithm dependent, since other tractography methods or atlas choices would possibly result in different tractograms ^14^. In addition, the BMRC motor status does not necessarily detect subtle or apractic motor deficits which might correlate with early tumor effects on the WM. Indeed, the main limitation for our study is the relatively small sample size we could include to perform the ML analysis. Moreover, we binarized motor deficits due to low number of samples per class. This led to an inaccuracy and affected the performance of our ML models. In order to develop models performing multiclass classification which could consider differences in degree of motor power (MRC = 1, 2, 3, 4, 5), larger samples would be needed per class. Furthermore, the dMRI data used for this study consists of a clinical single-shell acquisition, which is not optimal for fiber density measurements due to the incomplete attenuation of apparent extra-axonal signal ^14, 57^.

## Conclusion

In this study, we analyzed dMRI-based metrics to assess microstructural WM changes in correlation with the motor status of patients with gliomas in the motor system. We successfully developed SVM models to predict motor deficits in a heterogenous multivariate data set. ADC, FA and RD were highly predictive dMRI metrics. Additionally, we showed that dMRI metrics are better predictors than demographic and clinical variables, such as, age, glioma grade and RMT ratio. Careful selection and testing of ML modelling is mandatory to prevent over- or underfitting and misinterpretation of data.

## Materials and Methods

### Patient cohort

We included 116 left- and right-handed adult patients in this study (43 females, 73 males, average age = 48.24 ± 16.47, age range 20-78). Only patients with an initial diagnosis of supratentorial, unilateral WHO grade II, III & IV gliomas (16 WHO grade II, 23 WHO grade III, 77 WHO grade IV) were included (*Table 1*). All tumors were infiltrating or immediately adjacent to M1 and/or the CST either in the left or right hemisphere. Patients with recurrent tumors, previous radio-chemotherapy or multilocular tumors were not included. The motor status was graded preoperatively according to the Medical Research Council (MRC) scale for muscle power. Grade, 0 means no muscle power and 5 means full muscle strength. All patients with MRC<5 were assigned to the group with motor deficits (class 1), others (MRC = 5) were assigned to the group without motor deficits (class 0).

### Image acquisition

MRI data were acquired preoperatively at Charité University Hospital, Berlin, Department of Neuroradiology. The center performed scans on 3T Siemens Skyra scanner with dedicated 32-channel head/neck high count coil. The protocol included whole brain high-resolution structural data, contrast enhanced T1-weighted images, with TR/TE/TI 2300/2.32/900 ms flip angle = 9°, field of view (FOV) = 256 × 256, 192 sagittal slices, 1mm isotropic resolution, acquisition time: 5 min as well as a single shell diffusion weighted volume with TR/TE 7500/95ms, 2×2×2 mm^3^ voxels, 128 × 128 matrix, 60 axial slices, with 40 equally distributed orientations for diffusion-sensitizing gradients at b-value of 1000 s/mm^2^, for a total acquisition time of 12 minutes.

### Transcranial Magnetic Stimulation (TMS)

Non-invasive functional motor mapping of both ipsilesional and contralesional hemispheres was performed in each patient using navigated transcranial magnetic stimulation (nTMS) with NeXimia Navigated Brain Stimulation (Nexstim Oy, Helsinki, Finland). Each patient’s head was registered to the structural MRI and the composite muscle action potentials were captured by the integrated electromyography unit (EMG) (sampling rate 3 kHz, resolution 0.3 mV; Neuroline 720, Ambu). The muscle activity (motor evoked potential, MEP amplitude ≥ 50 μV) was recorded by surface electrodes on the abductor pollicis brevis and first dorsal interosseous. Initially, the first dorsal interosseous hotspot, defined as the stimulation area that evoked the strongest MEP, was determined. Subsequently, the resting motor threshold, defined as the lowest stimulation intensity that repeatedly elicits MEPs, was defined using a threshold-hunting algorithm within the Nexstim eximia software. Mapping was performed at 105% resting motor threshold and 0.25 Hz. All MEP amplitudes > 50 μV (peak to peak) were considered as motor positive responses and exported in the definitive mapping ^58^. The subject-specific positive responses of the first dorsal interosseous were exported as binary 3 mm^3^ voxel masks per response in the T1 image space.

### Preprocessing and processing of MRI data

Preprocessing and processing of MRI data was performed as described earlier ^14^. Briefly, all T1 images were linearly (affine) registered to the dMRI data sets using Advanced Normalization Tools (ANTs) ^59, 60^. Furthermore, we registered the human motor area template (HMAT) atlas to subject space with ANTs using the Symmetric Normalization (SyN) transformation model ^59, 61^ to obtain M1 seeding ROIs ^59, 61^. The preprocessing of dMRI data included the following and was performed within MRtrix3 ^56^ in sequential order: denoising ^62^, removal of Gibbs ringing artefacts ^63^, correction of subject motion ^64^, eddy-currents ^65^, and susceptibility-induced distortions ^66^ in FSL ^67^, and subsequent bias field correction with ANTs N4 ^68^. Each dMRI data set and processing step was visually inspected for outliers and artifacts. Scans with excessive motion were initially excluded based on a predefined threshold (if > 10% outlier slices, however this was not the case in the current cohort). We upsampled the dMRI data to a 1.3 mm isotropic voxel size before computing FODs to increase anatomical contrast and improve downstream tractography results and statistics ^69^. To obtain AD, ADC, FA, RD scalar maps, we first used diffusion tensor estimation using iteratively reweighted linear least squares estimator, resulting in scalar maps of tensor-derived parameters ^2,70^ For voxel-wise modelling we used a robust and fully automated and unsupervised method. This method allowed to obtain 3-tissue response functions for white and gray matter and cerebrospinal fluid (CSF) from our data with the use of spherical deconvolution for subsequent usage in multi-tissue CSD-based tractography ^25, 28, 71^.

### Tractography

Probabilistic multi-tissue tractography was performed based on the white matter FODs with the iFOD2 algorithm ^72^ as described earlier ^14^, with the slight modification of using the above mentioned HMAT atlas derived M1 seeding ROI ^73^. In brief, an inclusion ROI was defined in the medulla oblongata, tracking parameters were set to default with an FOD amplitude cutoff-value of 0.1, a streamline minimum length of 5 × voxel size and a maximum streamline length of 100 × voxel size. For each CST tractogram, we computed 5000 streamlines per hemisphere. Each streamline per tractogram was resampled along its length to 100 equidistant points. Subsequently, we mapped AD, ADC, FA, FD, and RD scalar metrics along the derived 100 equidistant points per streamline.

### Data Preparation

We generated dMRI-based CST profiles, by which AD, ADC, FA, FD, and RD were quantified at 100 segments along the CST using the values of the 100 points per streamlines. To create a tract profile that is robust to outliers, we used two different methods and compared the results. In the first method, we calculated the median (Mdn) values across the 5000 streamlines per tractogram along its 100 segments. In the second method, we computed the segment-wise weighted mean (M) of the dMRI measures across streamlines. The streamline-wise contribution was weighted by the inverse Mahalanobis distance of the streamlines from the tract core (mean). Streamlines that were more distant from M were considered less important ^17^. We used both ipsi- and contralesional CST profiles as input features (predictor variables), thus, the dimension of the imaging-based feature space was 1000 (5 metrics × 2 hemispheres × 100 segments = 1000 features).

## Statistical analysis

Statistical analysis and data visualization were carried out using Python 3.8.6. The main packages used were Scipy ^74^, Seaborn ^75^, Statsmodels ^76^ and Matplotlib ^77^. To compare categorical variables, Fisher’s exact test (when the expected frequency was less than five per category) or Pearson’s chi-squared test (for larger values) were employed. Two-tailed Student’s *t*-tests or Mann–Whitney *U* tests (Wilcoxon rank-sum test) were performed to compare continuous variables. Effect size (r) for Mann–Whitney *U-*statistics was calculated as the *Z*-statistic divided by the square root of the number of samples. A significance level of *p* < .05 was considered as cutoff. With respect to multiple comparison analyses, statistically significant *p*-values were false discovery rate (FDR) corrected using the Benjamini-Hochberg (BH) procedure ^78^. For all univariate statistical analyses Mdn-based tract profiles were used.

Canonical correlation analysis (CCA) was performed as a multivariate correlation analysis to identify and measure the association among all dMRI-based extracted features (both in ipsi- and contralesional hemispheres) and age ^79^. This analysis extracts meaningful information from a pair of data sets, dMRI-based features and age, by seeking pairs of linear combinations from two sets of variables with the maximum pairwise correlation. CCA was performed both on the patient’s cohort and on each patients’ group (class 0; class 1) separately. A more detailed analysis investigating the associations between each metrics and age was also performed.

### SVM classification

SVM has gained a widespread application in the neuroimaging context as either a classification or regression method ^80, 81^. The aim of an SVM classifier is to find hyperplanes with maximal margins between classes. As a supervised machine learning method, SVM can be extended to complex instances that are not linearly separable using so-called kernel tricks ^82, 83^. Kernel techniques map input features from one space to a higher dimensional feature space in which different classes can be distinguished by a separating hyperplane. All ML analysis methods should be balanced between their predictive accuracy and descriptive power ^84^. Accordingly, in the present study, we developed different models based SVM method using dMRI-based features, demographic and clinical variables to predict the motor status (class 0; class1) preoperatively, cf. *Fig. 5*.

**Figure 5:**
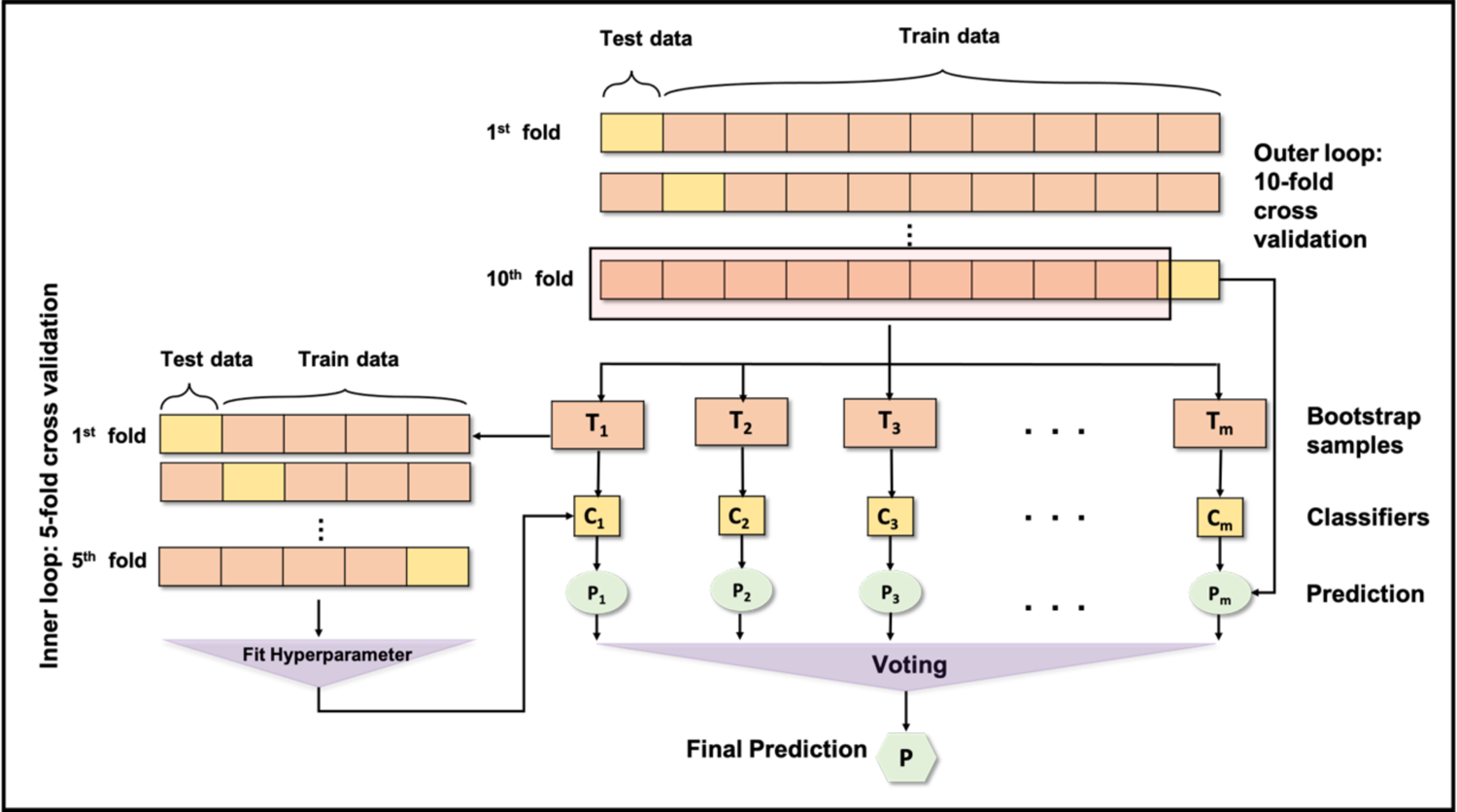
Visual summary of the machine learning pipeline with nested cross-validation and bootstrapping; First, test and train sets are selected per fold of outer loop CV. Next, numbers of (e.g., 1000) new synthetic datasets (T_i_) are generated from training set by randomly sampling from it. Hyper parameters of the classifiers (C_i_) are optimized within the inner loop; finally, estimator (P_i_) for each of the synthetic set is found and the result/prediction (P) are voted across them.

Some segments along tract profiles were missed (not-a-number value) when the tract profiles were generated. These missing values were imputed using two different interpolation methods ^85^, a) Mdn and b) k-nearest neighbor (KNN) ^86^. Before fitting a model to our data, imputation of missing values and standardization were performed. In order to enable our classifier to learn from low and high variance metrics, we removed each feature’s M and scaled it to a unit variance (z-score). Training and testing set within each cross validation were standardized separately by M and STD derived from the training set to prevent information leakage between testing and training data sets.

Four Different SVM models were trained and tested using Mdn-based and Mahalanobis-based weighted mean tract profiles with above-mentioned interpolation methods for imputation of missing values (**SVM_1**: Mdn-based imputation method and Mdn-based tract profile; **SVM_2**: KNN-based imputation method and Mdn-based tract profile; **SVM_3**: Mdn-based imputation methods and Mahalanobis-based weighted mean tract tract profile; **SVM_4**: KNN-based imputation method and Mahalanobis-based weighted mean tract profile).

As a preprocessing step, to reduce the high dimensional imaging-based feature space, a set of statistical features was calculated as a high-level representation to measure different properties of dMRI-based tract profiles’ distributions. We extracted M as a central tendency, STD as a measure of variability and kurtosis (KU) and skewness (SK) as measures of shape, *Fig. 6.* The tract profile statistics were calculated for ipsi- and contralesional tractograms (4 measures × 5 metrics × 2 hemispheres = 40 features) and were fed into the models. We further incorporated patients’ demographics and clinical data such as age, gender, tumor grade, tumor location and RMT ratio, and fed them into the aforementioned models and compared the results.

**Figure 6.**
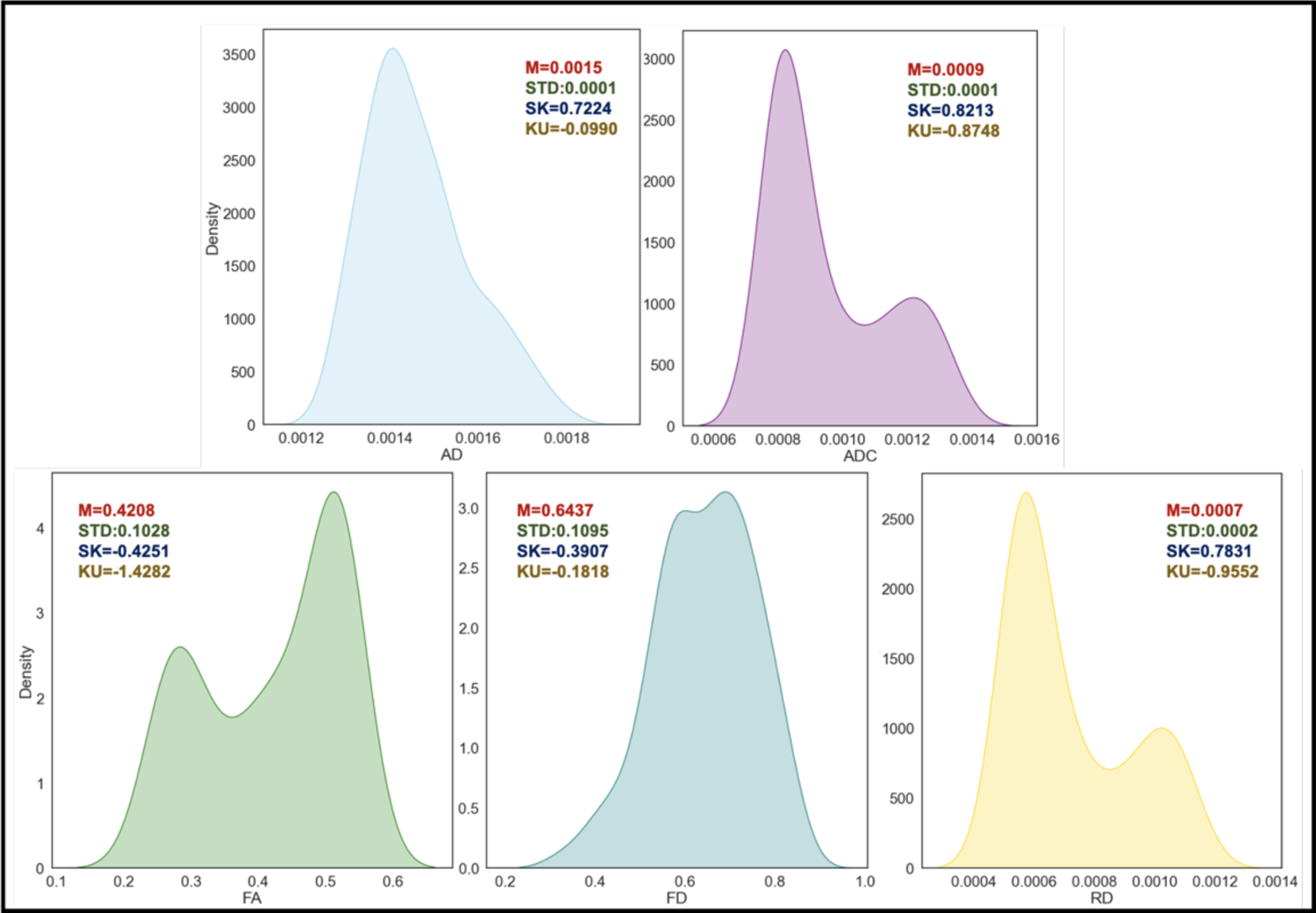
Kernel density estimation (KDE) plot for approximating the underlying probability density function for all dMRI-based ipsilesional tract profiles with the corresponding histogram-based features (4 features: M, STD, SK and KU) for a specific patient; a male patient (age:81) with preoperative motor deficits (class 1) and glioma WHO grade IV in the left hemisphere.

The most relevant features were selected using SVM-RFE ^37, 38^, which was intended to select subsets of features from the original rich feature space before doing the actual learning. This method recursively removes features that contribute least to the prediction based on the linear SVM classifier weight coefficients. Subsequently, selected features were used to train and validate the SVM model with linear kernel.

To investigate how well each dMRI metric (e.g., AD, ADC, FA, FD, RD) performed in classifying the patients with respect to their motor status, different SVM models were trained and tested, e.g., SVM_AD, SVM_ADC, SVM_FA, SVM_FD and SVM_RD, with KNN-based imputation method for missing values and Mdn-based tract profile. To assess the predictive power of patients’ demographics and clinical variables regardless of imaging-based features (when ignoring the neuroimaging analysis pipeline), an SVM model (SVM_clinical) was developed using only patients’ age, tumor WHO grade, tumor location, gender and RMT.

Additionally, a model was developed using all values of ipsi- and contralesional dMRI-based tract profiles without performing above mentioned feature extraction method. To reduce the high-dimensional imaging-based feature space (1000), principal component analysis (PCA) ^39, 40^ was performed on MdN-based tract profiles and the first 4 components were fed into an SVM model with linear kernel (SVM_5) using KNN-based imputation method.

We evaluated our models (SVM_1-5; SVM_AD-RD; SVM_clinical) using nested CV with a 10-fold outer loop and a 5-fold inner loop. Our model key hyperparameters C for penalty ^87^ was optimized in the inner CV loop and the best performing model was applied to the outer CV loop test set to evaluate the model selected by the inner loop. C was tested from 0 to 5 and 0 to 10 with a 0.1 step-size, respectively.

Our dataset was imbalanced because the proportion of patients with motor deficits to patients without motor deficits was nearly 1 to 2 (cf. *Table 1*). We used a stratified 10-fold CV to ensure that class distributions in each data split matched the distribution in the complete training dataset. We additionally assigned the class weights, W_j_, (class 1: MRC<0; class 0: MRC=5) inversely proportional to their respective frequencies as 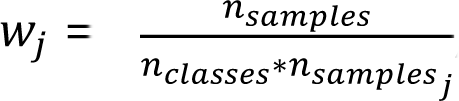, where n_samples_ is the number of samples, n_classes_ is the number of classes and n_samples_j is the number of samples per class.

Bootstrap aggregating (bagging) has been introduced as a method to reduce the variance of a given estimator ^88^. Bagging involves applying an estimator to multiple bootstrap samples and voting the results across them. These estimators can use CV themselves to select fine-tuning parameters trading off bias and variance of the bootstrap sample-specific candidate estimators. We used this approach in our models (SVM_1-4; SVM_AD-RD) with 1000 resampled training sets per fold of outer loop CV and lastly voted among all 1000 generated models.

We evaluated the performance of our models using the overall accuracy, the ratio of correctly predicted samples over the entire cohort, sensitivity, specificity and the area under the receiver operating curve (AUC).

## Data availability statement

Parts of the data that support the findings of this study are not publicly available due to information that could compromise the privacy of the research participants but are available from the corresponding author on reasonable request. However, code we have used is openly available on https://github.com/CUB-IGL/Machine-learning-based-prediction-of-motor-status-in-glioma-patients-using-dMRI-metrics-along-CST and is referred to at the corresponding passage in the article.

## Acknowledgements

BS, LSF and TP acknowledge the support of the Cluster of Excellence Matters of Activity. Image Space Material funded by the Deutsche Forschungsgemeinschaft (DFG, German Research Foundation) under Germany’s Excellence Strategy–EXC 2025 – 390648296. T.R. received support from the Finnish Cultural Foundation.

## Funding

Deutsche Forschungsgemeinschaft German Research Foundation (DFG) under Germany’s Excellence Strategy–EXC 2025 – 390648296.

## Author information

Affiliations

Cluster of Excellence Matters of Activity. Image Space Material, Humboldt Universität zu Berlin, Germany

**Charité – Universitätsmedizin Berlin, Department of Neurosurgery, Berlin, Germany**

Boshra Shams, Ziqian Wang, Peter Vajkoczy, Thomas Picht, Lucius S. Fekonja

Digital Health - Machine Learning, Hasso Plattner Institute, University of Potsdam Potsdam, Germany

**Hasso Plattner Institute for Digital Health, Icahn School of Medicine at Mount Sinai, New NY, USA**

Christoph Lippert

Department of Neuroscience and Biomedical Engineering, Aalto University School of Science, Espoo, Finland

Baran Aydogan, Timo Roine

Department of Psychiatry, Helsinki University and Helsinki University Hospital, Helsinki, Finland

Baran Aydogan

Turku Brain and Mind Center, University of Turku, Turku, Finland

Timo Roine

## Author contributions

Conceptualization: L.S.F., T.P. Methodology: L.S.F., B.S. Investigation: L.S.F., B.S. Visualization: L.S.F., B.S. Funding acquisition: L.S.F., T.P., P.V. Supervision: L.S.F., T.P., C.L., P.V., B.A., T.R. Writing – original draft: B.S., L.S.F., T.R., B.A., C.L., T.P. Corresponding authors Correspondence to Dr. Lucius S. Fekonja. Orcid:0000-0003-1973-4410

## Competing interests

The authors report no competing interests.

## Ethical standard

The study proposal is in accordance with ethical standards of the Declaration of Helsinki and was approved by the Ethics Commission of the Charité University Hospital (#EA1/016/19). All patients provided written informed consent for medical evaluations and treatments within the scope of the study.

## Supplementary information

**Supplementary Table 1:** CCA correlation coefficients

|  |  |
| --- | --- |
| CCX_1 | CCY_1 |
| 0.3686912166385593 | 1.0104415505824895 |
| 0.10362663900566732 | -0.15370543189728422 |
| 0.04403315110177353 | 0.45900350624996517 |
| 0.03699685340390663 | -0.5213307947856338 |
| -0.17735774359011147 | -0.5826016886003588 |
| 0.14144572832025423 | 0.8266288691383148 |
| -0.14389351484917856 | -0.8889561576739835 |
| 0.027427938022573206 | -0.7664143700445336 |
| 0.0484294883680349 | -0.398789007156184 |
| -0.048689189391995125 | 0.09137814336161552 |
| 0.04167285754217588 | 1.3780669134708392 |
| 0.2590717825660527 | 1.1942542320266645 |
| -0.1472697320691347 | -0.46005990097090893 |
| 0.07562702673713945 | 0.09137814336161552 |
| -0.039559172595529324 | 0.33646171862051527 |
| -0.06293601384047992 | 1.0717124443972146 |
| -0.2553732760525167 | -1.5016650958212328 |
| -0.14105888518587928 | -1.0727688391181582 |
| -0.0473670145207456 | -1.317852414377058 |
| -0.3988483653797505 | -1.7467486710801325 |
| -0.13117284178465785 | -0.7664143700445336 |
| -0.42049385794971866 | -1.0114979453034334 |
| 0.2361710754481009 | 1.1942542320266645 |
| 0.18752567882663085 | 1.439337807285564 |
| -0.006205673042270071 | 0.64281618769414 |
| 0.04854366866083257 | -0.21497632571200917 |
| -0.051634421737504155 | -0.398789007156184 |
| 0.006099809021180888 | 0.7653579753235898 |
| -0.012605023879954158 | 0.64281618769414 |
| 0.2444290368590513 | 1.500608701100289 |
| 0.3569216704829967 | 1.500608701100289 |
| -0.16985814362474877 | -0.2762472195267341 |
| 0.3716031027967968 | 1.3167960196561141 |
| -0.1691482163357569 | 0.2139199309910654 |
| -0.05928120851037519 | -1.440394202006508 |
| -0.19763593014147318 | -0.8889561576739835 |
| -0.25902187628122353 | -1.5016650958212328 |
| 0.13981394649063342 | -0.031163644267834356 |
| -0.23389167446150771 | -1.5629359896359578 |
| -0.25864717821041683 | -0.398789007156184 |
| -0.2377550400139979 | -1.1340397329328833 |
| -0.30359358883485676 | -1.5016650958212328 |
| -0.032678460551597066 | -1.8080195648948576 |
| 0.03372427650683013 | 0.15264903717634046 |
| -0.1035217616529111 | 0.8266288691383148 |
| -0.11259671233882773 | -0.9502270514887085 |
| -0.143517620363094 | -2.053103140153757 |
| -0.21655465058404733 | -1.5016650958212328 |
| 0.017365049436355508 | -0.031163644267834356 |
| 0.13399513399736385 | 0.7040870815088649 |
| 0.016772346545457117 | 0.64281618769414 |
| -0.08828321690555402 | -0.398789007156184 |
| 0.12267800447446522 | -0.33751811334145904 |
| -0.008023121242003661 | 0.45900350624996517 |
| 0.3081178290546914 | 0.03010724954689058 |
| -0.09312951597811121 | -0.15370543189728422 |
| 0.09283923334000155 | 0.33646171862051527 |
| -0.15162597058252636 | -0.5826016886003588 |
| -0.16581789182903844 | 1.1329833382119394 |
| -0.413272748943199 | -1.7467486710801325 |
| -0.2971687550920949 | -1.317852414377058 |
| -0.2928975011682272 | -0.398789007156184 |
| -0.23481293758078048 | -0.46005990097090893 |
| 0.14615638714669485 | -0.09243453808255929 |
| 0.047844373811946506 | 0.7040870815088649 |
| -0.1883070682285135 | 0.2751908248057903 |
| -0.07081406263180283 | -1.440394202006508 |
| -0.21716385487415982 | -1.317852414377058 |
| -0.36461738008333017 | -1.5629359896359578 |
| 0.18697497053090434 | 1.6844213825444638 |
| -0.09963706492817292 | -0.2762472195267341 |
| -0.13256894516837506 | 0.09137814336161552 |
| 0.06837227386648927 | 0.15264903717634046 |
| 0.46042944838314914 | 1.6844213825444638 |
| 0.076918596282874 | 0.7653579753235898 |
| 0.08497298065577868 | 0.33646171862051527 |
| 0.28343681857808045 | 1.0104415505824895 |
| 0.1475016935762437 | 0.7653579753235898 |
| 0.21206897900667582 | 0.9491706567677647 |
| -0.06116526778040114 | 0.7653579753235898 |
| 0.24563959901547888 | 1.439337807285564 |
| -0.026158822161424622 | 0.7040870815088649 |
| -0.002682222031399069 | -0.5826016886003588 |
| 0.16262747409970063 | 0.7040870815088649 |
| 0.27634393941922125 | 0.64281618769414 |
| 0.2947311692851994 | -0.21497632571200917 |
| 0.26758830157586366 | 0.8266288691383148 |
| -0.3692862370625389 | -1.8080195648948576 |
| 0.2964645250518494 | -0.46005990097090893 |
| 0.048033742165981774 | -0.8889561576739835 |
| 0.021088377936743785 | 0.7653579753235898 |
| 0.12823268308729202 | 1.3780669134708392 |
| 0.07055791424051508 | -0.5826016886003588 |
| 0.15975435631499993 | -0.33751811334145904 |
| -0.08736568282474619 | -0.2762472195267341 |
| -0.06829483834326558 | -1.6242068834506826 |
| 0.008068885726550568 | 0.5202744000646901 |
| 0.0890820866674716 | -0.7664143700445336 |
| 0.22251321030151677 | 1.1329833382119394 |
| -0.17171582364615545 | -1.317852414377058 |
| -0.45049276407935457 | -1.1953106267476081 |
| 0.15036583286041327 | 1.3167960196561141 |
| 0.01902318444759854 | -0.031163644267834356 |
| 0.07770181943201215 | 0.45900350624996517 |
| 0.08816528027367487 | -0.15370543189728422 |
| 0.11689277973220724 | 1.8682340639886388 |
| 0.10109922838249685 | 0.03010724954689058 |
| 0.040047488629599615 | 0.9491706567677647 |
| 0.1029032984760647 | 1.1329833382119394 |
| -0.25296601879859976 | 0.03010724954689058 |
| -0.02141469372431091 | 0.09137814336161552 |
| 0.06388993725933284 | 0.2751908248057903 |
| -0.05071430452563272 | -1.1953106267476081 |
| 0.09693971124921125 | 1.561879594915014 |
| 0.06023449221208867 | 2.0520467454328135 |
| 0.5762751146592526 | 1.3780669134708392 |

**Supplementary Table 2:** Corrected *p-*values (FDR-BH) along ipsilesional CST profiles

| AD | ADC | FA | FD | RD |
| --- | --- | --- | --- | --- |
| 0.011136856046762069 | 0.011462777718257914 | 0.27965843978691385 | 0.336694391190237 | 0.023952035688187028 |
| 0.016744609325347045 | 0.011462777718257914 | 0.8614222560019962 | 0.8274002614710084 | 0.02009184498865534 |
| 0.03015882120803534 | 0.017049460927474815 | 0.7461304467136712 | 0.9141143985149782 | 0.023928278183991784 |
| 0.03463002505999671 | 0.017049460927474815 | 0.4992714530269339 | 0.9141143985149782 | 0.02000370924149419 |
| 0.03463002505999671 | 0.01609853273759804 | 0.33362888576119026 | 0.9888779233376925 | 0.01934585148806892 |
| 0.02929096906969759 | 0.015070328030297207 | 0.2411192776556674 | 0.9982066951663279 | 0.02000370924149419 |
| 0.02929096906969759 | 0.016414059129873117 | 0.1882602269445827 | 0.9982066951663279 | 0.02301654840936339 |
| 0.03171839305313943 | 0.01609853273759804 | 0.16357062674222658 | 0.9627634760330447 | 0.023377814261011088 |
| 0.025874570423586082 | 0.011767707372551789 | 0.14071099944515375 | 0.9141143985149782 | 0.02026124322500367 |
| 0.02121149154014225 | 0.011462777718257914 | 0.13618760908001423 | 0.9141143985149782 | 0.018164815368189098 |
| 0.021243206088001957 | 0.011462777718257914 | 0.137593259106739 | 0.9539734530140437 | 0.018164815368189098 |
| 0.02282886248425515 | 0.011767707372551789 | 0.14071099944515375 | 0.9627634760330447 | 0.01890593775097565 |
| 0.02282886248425515 | 0.011767707372551789 | 0.16357062674222658 | 0.9732034434130821 | 0.02009184498865534 |
| 0.026836322510219602 | 0.011767707372551789 | 0.14071099944515375 | 0.9542175652766098 | 0.01890593775097565 |
| 0.03171839305313943 | 0.011767707372551789 | 0.13618760908001423 | 0.9542175652766098 | 0.018164815368189098 |
| 0.03171839305313943 | 0.011767707372551789 | 0.14071099944515375 | 0.9141143985149782 | 0.01871782297626124 |
| 0.03463002505999671 | 0.011462777718257914 | 0.13618760908001423 | 0.7993414709537926 | 0.018164815368189098 |
| 0.055242742382381436 | 0.01176252940160753 | 0.1123439735986235 | 0.7735097655761669 | 0.018164815368189098 |
| 0.06098530177861444 | 0.01637697785263732 | 0.12522090303602273 | 0.732829340561692 | 0.02049227013218372 |
| 0.056990371311264 | 0.02023806896489688 | 0.14379811183280242 | 0.670781882771652 | 0.023377814261011088 |
| 0.055242742382381436 | 0.019547520296756882 | 0.17010815741260482 | 0.6256598717427512 | 0.024336388411057005 |
| 0.06098530177861444 | 0.02323681379112752 | 0.21170746187000897 | 0.6042199620338617 | 0.029665195952593204 |
| 0.08994367978888763 | 0.0338046226196035 | 0.2943903786876607 | 0.6042199620338617 | 0.04359606908955554 |
| 0.12999433043589012 | 0.04208287858689789 | 0.3982503178400343 | 0.6042199620338617 | 0.06286515888821723 |
| 0.14699471121546623 | 0.03634301769573582 | 0.5425887833684939 | 0.6042199620338617 | 0.07156173355217792 |
| 0.15608847981541277 | 0.028789716935267145 | 0.6291283969128854 | 0.5822955665604527 | 0.06969162288672615 |
| 0.16651563180443216 | 0.026615777124084465 | 0.6012948757163232 | 0.4325106201914536 | 0.06691954796741748 |
| 0.17812251369044055 | 0.02917542685817119 | 0.5734108400585972 | 0.2819084518363021 | 0.06883391740536415 |
| 0.19923711247861567 | 0.02917542685817119 | 0.5326023926399954 | 0.19195509787067483 | 0.061758018317453554 |
| 0.2266382440708272 | 0.02832680226944813 | 0.4613677639298348 | 0.1485938230016407 | 0.04440187444481049 |
| 0.2266382440708272 | 0.023864080709273018 | 0.3982503178400343 | 0.1485938230016407 | 0.03468365486650289 |
| 0.21269741546085077 | 0.024831482722872774 | 0.38007842641007183 | 0.1485938230016407 | 0.03468365486650289 |
| 0.20946354884691123 | 0.027700280201350142 | 0.3896753234923075 | 0.1485938230016407 | 0.040900643477244016 |
| 0.17812251369044055 | 0.025011095415107754 | 0.38563100423617064 | 0.1485938230016407 | 0.04083623304439021 |
| 0.13581417513481156 | 0.018157248529293974 | 0.31670973161260846 | 0.1485938230016407 | 0.03291789003938622 |
| 0.11889043648836928 | 0.015956387876704864 | 0.2786671226853675 | 0.1485938230016407 | 0.025887432625942456 |
| 0.10458775476628153 | 0.012048039939190944 | 0.213333119674164 | 0.1485938230016407 | 0.023377814261011088 |
| 0.0795752340698011 | 0.011462777718257914 | 0.14492614860552588 | 0.1485938230016407 | 0.01934585148806892 |
| 0.05863356146822964 | 0.011462777718257914 | 0.1123439735986235 | 0.1485938230016407 | 0.018164815368189098 |
| 0.056990371311264 | 0.011462777718257914 | 0.07994206707289495 | 0.1485938230016407 | 0.018076665667520586 |
| 0.05792775878183194 | 0.011462777718257914 | 0.05959326596819153 | 0.1485938230016407 | 0.017421757849957253 |
| 0.05401262186603804 | 0.011462777718257914 | 0.04550526922929115 | 0.1485938230016407 | 0.01588723194494714 |
| 0.04470889004892224 | 0.011462777718257914 | 0.04078425140010971 | 0.1485938230016407 | 0.01588723194494714 |
| 0.04470889004892224 | 0.011462777718257914 | 0.04078425140010971 | 0.1485938230016407 | 0.01588723194494714 |
| 0.04870401647324613 | 0.011462777718257914 | 0.04078425140010971 | 0.1485938230016407 | 0.017421757849957253 |
| 0.05863356146822964 | 0.011497714725035172 | 0.04078425140010971 | 0.1485938230016407 | 0.018164815368189098 |
| 0.07901994313636655 | 0.011767707372551789 | 0.04078425140010971 | 0.15226213864144128 | 0.018164815368189098 |
| 0.09306233751815475 | 0.012747849113972198 | 0.041863934899368835 | 0.17571266360674254 | 0.018164815368189098 |
| 0.10458775476628153 | 0.013619963168304565 | 0.04550526922929115 | 0.18890654634936266 | 0.018164815368189098 |
| 0.11777182152502443 | 0.014565874125948888 | 0.04740731515010825 | 0.18907617870645227 | 0.01890593775097565 |
| 0.11889043648836928 | 0.01609853273759804 | 0.05137638799979671 | 0.18907617870645227 | 0.02000370924149419 |
| 0.12078500583653652 | 0.017999557623633866 | 0.061543138659016054 | 0.19233816913746687 | 0.021779460615798985 |
| 0.11889043648836928 | 0.021012431943190846 | 0.07780416004578886 | 0.23259677308874643 | 0.023928278183991784 |
| 0.10458775476628153 | 0.023614098401770214 | 0.10144973516677312 | 0.27743331853289555 | 0.026462791433194653 |
| 0.07952113868114824 | 0.024431221390329605 | 0.12522090303602273 | 0.336694391190237 | 0.03157076588168193 |
| 0.05863356146822964 | 0.02323681379112752 | 0.14071099944515375 | 0.43735071883222243 | 0.03550853313730138 |
| 0.04773526620930219 | 0.02171390340272644 | 0.16654117355485182 | 0.478215731155554 | 0.0381613645019177 |
| 0.03463002505999671 | 0.017893599449023478 | 0.20009489545153625 | 0.6042199620338617 | 0.0381613645019177 |
| 0.024382381326636202 | 0.015070328030297207 | 0.22699672556737807 | 0.6859842391892909 | 0.03588214425467024 |
| 0.016744609325347045 | 0.011894922454589696 | 0.2411192776556674 | 0.7695288387565576 | 0.03313585756572504 |
| 0.01392081883561952 | 0.011497714725035172 | 0.2411192776556674 | 0.789757454540459 | 0.02851365755958812 |
| 0.011136856046762069 | 0.011462777718257914 | 0.2411192776556674 | 0.789757454540459 | 0.02566864216123481 |
| 0.011136856046762069 | 0.011462777718257914 | 0.2411192776556674 | 0.789757454540459 | 0.023928278183991784 |
| 0.011136856046762069 | 0.011462777718257914 | 0.2411192776556674 | 0.789757454540459 | 0.023278674419693245 |
| 0.011136856046762069 | 0.011462777718257914 | 0.2411192776556674 | 0.789757454540459 | 0.02301654840936339 |
| 0.011136856046762069 | 0.011462777718257914 | 0.25535977736224225 | 0.789757454540459 | 0.023278674419693245 |
| 0.011136856046762069 | 0.011462777718257914 | 0.2639165492491231 | 0.789757454540459 | 0.023928278183991784 |
| 0.011136856046762069 | 0.011462777718257914 | 0.2786671226853675 | 0.789757454540459 | 0.023952035688187028 |
| 0.011136856046762069 | 0.011462777718257914 | 0.27965843978691385 | 0.789757454540459 | 0.023952035688187028 |
| 0.011136856046762069 | 0.011462777718257914 | 0.2943903786876607 | 0.789757454540459 | 0.024732373214574433 |
| 0.011206775544242457 | 0.011462777718257914 | 0.31064768769730416 | 0.789757454540459 | 0.02518303307330361 |
| 0.01286524468623974 | 0.011462777718257914 | 0.31285805946128703 | 0.789757454540459 | 0.023952035688187028 |
| 0.014109536187488588 | 0.011462777718257914 | 0.30917252479607626 | 0.789757454540459 | 0.023928278183991784 |
| 0.014890466596799495 | 0.011462777718257914 | 0.27965843978691385 | 0.789757454540459 | 0.023278674419693245 |
| 0.018680309910050114 | 0.011462777718257914 | 0.25535977736224225 | 0.789757454540459 | 0.021394064999468276 |
| 0.02282886248425515 | 0.011462777718257914 | 0.22699672556737807 | 0.789757454540459 | 0.02000370924149419 |
| 0.02929096906969759 | 0.011462777718257914 | 0.16357062674222658 | 0.7695288387565576 | 0.019294243701493925 |
| 0.03463002505999671 | 0.011462777718257914 | 0.12522090303602273 | 0.6042199620338617 | 0.018164815368189098 |
| 0.04021560781878949 | 0.011497714725035172 | 0.07780416004578886 | 0.4658812642859192 | 0.018164815368189098 |
| 0.04470889004892224 | 0.011497714725035172 | 0.05088356958280326 | 0.3533492096113398 | 0.018164815368189098 |
| 0.04870401647324613 | 0.011497714725035172 | 0.04078425140010971 | 0.2770301593694052 | 0.018164815368189098 |
| 0.04870401647324613 | 0.011462777718257914 | 0.03245043014523583 | 0.23349455839247987 | 0.018033289115355795 |
| 0.04870401647324613 | 0.011462777718257914 | 0.02511995941577425 | 0.19195509787067483 | 0.01588723194494714 |
| 0.04470889004892224 | 0.011462777718257914 | 0.020169973894517256 | 0.17571266360674254 | 0.015527430091551199 |
| 0.043107132670720964 | 0.011462777718257914 | 0.018137062558646628 | 0.15226213864144128 | 0.013917474296056631 |
| 0.03463002505999671 | 0.011462777718257914 | 0.01662788252002256 | 0.15074839122312966 | 0.013323800502485033 |
| 0.03155622017843677 | 0.011462777718257914 | 0.01662788252002256 | 0.1485938230016407 | 0.012202570484895169 |
| 0.025574079303818092 | 0.011462777718257914 | 0.01662788252002256 | 0.1485938230016407 | 0.012202570484895169 |
| 0.022622328713007275 | 0.011462777718257914 | 0.01662788252002256 | 0.1485938230016407 | 0.012202570484895169 |
| 0.019106282780761756 | 0.011462777718257914 | 0.01662788252002256 | 0.1485938230016407 | 0.012202570484895169 |
| 0.016744609325347045 | 0.011462777718257914 | 0.01662788252002256 | 0.15226213864144128 | 0.012202570484895169 |
| 0.014890466596799495 | 0.011462777718257914 | 0.01662788252002256 | 0.18890654634936266 | 0.012202570484895169 |
| 0.014890466596799495 | 0.011462777718257914 | 0.018137062558646628 | 0.23349455839247987 | 0.012202570484895169 |
| 0.01393676144721457 | 0.011462777718257914 | 0.02424750538883141 | 0.3317422608885178 | 0.012202570484895169 |
| 0.011882195455281816 | 0.011462777718257914 | 0.035301485609796034 | 0.47676095184626227 | 0.012202570484895169 |
| 0.011136856046762069 | 0.011462777718257914 | 0.04078425140010971 | 0.6473393205321074 | 0.012202570484895169 |
| 0.011136856046762069 | 0.011462777718257914 | 0.06075201010117749 | 0.789757454540459 | 0.012202570484895169 |
| 0.011136856046762069 | 0.011462777718257914 | 0.10144973516677312 | 0.8871531125264345 | 0.012202570484895169 |
| 0.011136856046762069 | 0.011462777718257914 | 0.14492614860552588 | 0.9627634760330447 | 0.012202570484895169 |
| 0.011136856046762069 | 0.011462777718257914 | 0.6906425930218448 | 0.789757454540459 | 0.018164815368189098 |

**Supplementary Table 3:** Corrected *p*-values (FDR-BH) along the differences between ipsi- and contralesional CST profiles

| AD | ADC | FA | FD | RD |
| --- | --- | --- | --- | --- |
| 0.946710846397834 | 0.7807702814114579 | 0.37323011750235585 | 0.9775738965452768 | 0.5838802437081158 |
| 0.4818589402602446 | 0.556726936781187 | 0.8410825003874249 | 0.9040061752791977 | 0.5928992668593196 |
| 0.42558388377505574 | 0.29474932132860976 | 0.7166517983490064 | 0.8313145176230737 | 0.2686273630726987 |
| 0.3645473342101959 | 0.13266042076113374 | 0.3153792871266046 | 0.837624646639151 | 0.10380438744659091 |
| 0.3645473342101959 | 0.08866269318085154 | 0.18350084927364785 | 0.6495610369980465 | 0.07366599053088398 |
| 0.3735489881783121 | 0.1040775601787513 | 0.14168057428042644 | 0.6427869803426212 | 0.07634300862113903 |
| 0.4538768865847612 | 0.14407507449662693 | 0.11596421370663372 | 0.6328905221979035 | 0.10682710846045199 |
| 0.6588512122768341 | 0.1758508019417226 | 0.08669312988369528 | 0.6328905221979035 | 0.1278380980062686 |
| 0.751280416125299 | 0.21034427748509554 | 0.06482408763999176 | 0.6044532261016083 | 0.1278380980062686 |
| 0.7813800699684113 | 0.2979382908536028 | 0.06721042042700122 | 0.6328905221979035 | 0.15082288891412649 |
| 0.9034658406277295 | 0.6142184862401171 | 0.08669312988369528 | 0.6524646842379762 | 0.2686273630726987 |
| 0.8780239686427145 | 0.8968498784656392 | 0.12974703468049267 | 0.7553173382781562 | 0.4476555411542437 |
| 0.8469400277391879 | 0.9910317149864964 | 0.1553741744955487 | 0.8172452181173122 | 0.5291047581762206 |
| 0.7813800699684113 | 0.9602345364417547 | 0.15083470128058205 | 0.8155503794384331 | 0.5110852133591515 |
| 0.7813800699684113 | 0.9399810614796142 | 0.1553741744955487 | 0.9734637368410731 | 0.36904925261372795 |
| 0.8780239686427145 | 0.8788745710258984 | 0.19557810828249525 | 0.9775738965452768 | 0.3635930458206528 |
| 0.8780239686427145 | 0.756776441358386 | 0.2019855719955663 | 0.9734637368410731 | 0.3257185261513045 |
| 0.8780239686427145 | 0.6558705945376152 | 0.17995604433123588 | 0.9734637368410731 | 0.25840855408827007 |
| 0.9815595076030244 | 0.5975384727625117 | 0.18355486471250076 | 0.9734637368410731 | 0.26545028553115174 |
| 0.8780239686427145 | 0.5152732922843374 | 0.19322221671857648 | 0.9638786823435999 | 0.2402165384885349 |
| 0.8469400277391879 | 0.3735900802009081 | 0.18350084927364785 | 0.946826799458898 | 0.2010606427452123 |
| 0.8382341274883265 | 0.3368503135066111 | 0.17116087929412843 | 0.8809348111747528 | 0.1839953263949054 |
| 0.8469400277391879 | 0.4290329175270216 | 0.23067527627375958 | 0.9040061752791977 | 0.21890095410974983 |
| 0.875295214773319 | 0.4590821724197558 | 0.3120567807889906 | 0.9638786823435999 | 0.2402165384885349 |
| 0.7813800699684113 | 0.3533166992981862 | 0.3612151694221496 | 0.9899872919283637 | 0.21854516509838162 |
| 0.7388258758422597 | 0.21034427748509554 | 0.3203015998138497 | 0.9775738965452768 | 0.16149468799944056 |
| 0.6570449621119576 | 0.14407507449662693 | 0.22760500025472905 | 0.9495976721906235 | 0.1279680535406273 |
| 0.5275718685377806 | 0.1366278639994293 | 0.19761984533463753 | 0.7630581559197008 | 0.13944104288251719 |
| 0.4705315369348145 | 0.1131651695996264 | 0.18355486471250076 | 0.6704805735342625 | 0.1278380980062686 |
| 0.5030343148422163 | 0.09569297073289015 | 0.1553741744955487 | 0.6458222576050873 | 0.09361981768721692 |
| 0.4705315369348145 | 0.0831925205126628 | 0.1553741744955487 | 0.6458222576050873 | 0.08575717647527047 |
| 0.4538768865847612 | 0.09001828786424906 | 0.18350084927364785 | 0.643190672951719 | 0.09594313955215965 |
| 0.4538768865847612 | 0.1131651695996264 | 0.22945409490469965 | 0.6524646842379762 | 0.1278380980062686 |
| 0.4538768865847612 | 0.1131651695996264 | 0.2539560632242701 | 0.6704805735342625 | 0.1278380980062686 |
| 0.42558388377505574 | 0.08949225398151685 | 0.2019855719955663 | 0.6704805735342625 | 0.10982183469040709 |
| 0.4466102658776658 | 0.08295252674525776 | 0.18355486471250076 | 0.6795303498327848 | 0.09594533336998949 |
| 0.43468635751057527 | 0.08007254592202284 | 0.1553741744955487 | 0.6704805735342625 | 0.09309908885774293 |
| 0.42558388377505574 | 0.07085636337170394 | 0.13803205013157507 | 0.6524646842379762 | 0.07533978150946301 |
| 0.4168535932962531 | 0.07085636337170394 | 0.09511219401924051 | 0.6458222576050873 | 0.053407816047747116 |
| 0.42558388377505574 | 0.07085636337170394 | 0.0721607634931837 | 0.6427869803426212 | 0.052370196524349044 |
| 0.4538768865847612 | 0.07085636337170394 | 0.06482408763999176 | 0.6427869803426212 | 0.052370196524349044 |
| 0.4705315369348145 | 0.07085636337170394 | 0.053400683202913836 | 0.6328905221979035 | 0.052370196524349044 |
| 0.4818589402602446 | 0.07085636337170394 | 0.053400683202913836 | 0.6328905221979035 | 0.052370196524349044 |
| 0.5071634137125404 | 0.07085636337170394 | 0.053400683202913836 | 0.6044532261016083 | 0.052370196524349044 |
| 0.514652407761254 | 0.07582894780604216 | 0.053400683202913836 | 0.6044532261016083 | 0.052370196524349044 |
| 0.6024072846192386 | 0.07901464834480984 | 0.053400683202913836 | 0.6044532261016083 | 0.052370196524349044 |
| 0.7194119106828601 | 0.08095735925122848 | 0.053400683202913836 | 0.6044532261016083 | 0.052370196524349044 |
| 0.7813800699684113 | 0.08125374404500925 | 0.053400683202913836 | 0.6044532261016083 | 0.052370196524349044 |
| 0.8469400277391879 | 0.08125374404500925 | 0.053400683202913836 | 0.6044532261016083 | 0.052370196524349044 |
| 0.8780239686427145 | 0.08125374404500925 | 0.053400683202913836 | 0.6044532261016083 | 0.052370196524349044 |
| 0.8780239686427145 | 0.0836934139524262 | 0.053400683202913836 | 0.6044532261016083 | 0.053407816047747116 |
| 0.8780239686427145 | 0.09116335398184212 | 0.053400683202913836 | 0.6044532261016083 | 0.05556674288447302 |
| 0.8780239686427145 | 0.10100102297715273 | 0.05533487826930164 | 0.6044532261016083 | 0.06250788708720481 |
| 0.875295214773319 | 0.1131651695996264 | 0.06482408763999176 | 0.6328905221979035 | 0.07239593478892002 |
| 0.7813800699684113 | 0.1131651695996264 | 0.08409221559866405 | 0.6328905221979035 | 0.08177527558658071 |
| 0.6570449621119576 | 0.10928301473566718 | 0.09717724750201813 | 0.6427869803426212 | 0.09086239954748633 |
| 0.514652407761254 | 0.09808096357351669 | 0.11796322308970694 | 0.6427869803426212 | 0.09175029282610467 |
| 0.464352857656369 | 0.08295252674525776 | 0.13715506819861734 | 0.6458222576050873 | 0.09105716260312426 |
| 0.42558388377505574 | 0.07644946202713564 | 0.14318687193700194 | 0.6495610369980465 | 0.08433823700610824 |
| 0.3645473342101959 | 0.07085636337170394 | 0.15083470128058205 | 0.6524646842379762 | 0.07555886579618676 |
| 0.35729119733545944 | 0.07085636337170394 | 0.15083470128058205 | 0.6704805735342625 | 0.07016266499101857 |
| 0.34112341487223097 | 0.07085636337170394 | 0.15083470128058205 | 0.6524646842379762 | 0.06456733653832186 |
| 0.34112341487223097 | 0.07085636337170394 | 0.15083470128058205 | 0.6495610369980465 | 0.0602467574325961 |
| 0.3538345288232321 | 0.07085636337170394 | 0.14318687193700194 | 0.6427869803426212 | 0.05518072776899828 |
| 0.3585798533491954 | 0.07085636337170394 | 0.12725758000572143 | 0.6427869803426212 | 0.053407816047747116 |
| 0.3585798533491954 | 0.07085636337170394 | 0.11596421370663372 | 0.6421060214145352 | 0.053407816047747116 |
| 0.36262055396206444 | 0.07085636337170394 | 0.10220940273130472 | 0.6328905221979035 | 0.053407816047747116 |
| 0.36262055396206444 | 0.07085636337170394 | 0.09717724750201813 | 0.6328905221979035 | 0.053407816047747116 |
| 0.3645473342101959 | 0.07085636337170394 | 0.09717724750201813 | 0.6328905221979035 | 0.053407816047747116 |
| 0.3645473342101959 | 0.07085636337170394 | 0.09360098287455311 | 0.6328905221979035 | 0.053407816047747116 |
| 0.3645473342101959 | 0.07085636337170394 | 0.0887404727518205 | 0.6427869803426212 | 0.053407816047747116 |
| 0.3735489881783121 | 0.07085636337170394 | 0.08366176387172798 | 0.6427869803426212 | 0.052370196524349044 |
| 0.4194614171345507 | 0.07085636337170394 | 0.07711797313590166 | 0.6524646842379762 | 0.052370196524349044 |
| 0.42558388377505574 | 0.07085636337170394 | 0.06721042042700122 | 0.6704805735342625 | 0.052370196524349044 |
| 0.4538768865847612 | 0.07085636337170394 | 0.06482408763999176 | 0.7553173382781562 | 0.052370196524349044 |
| 0.4538768865847612 | 0.07085636337170394 | 0.06482408763999176 | 0.7630581559197008 | 0.052370196524349044 |
| 0.464352857656369 | 0.07085636337170394 | 0.06482408763999176 | 0.7553173382781562 | 0.052370196524349044 |
| 0.4700769606431286 | 0.07582894780604216 | 0.05533487826930164 | 0.6704805735342625 | 0.052370196524349044 |
| 0.4705315369348145 | 0.07644946202713564 | 0.053400683202913836 | 0.6524646842379762 | 0.052370196524349044 |
| 0.464352857656369 | 0.07644946202713564 | 0.053400683202913836 | 0.6495610369980465 | 0.052370196524349044 |
| 0.464352857656369 | 0.07644946202713564 | 0.053400683202913836 | 0.6458222576050873 | 0.052370196524349044 |
| 0.4538768865847612 | 0.0763728527061631 | 0.053400683202913836 | 0.6458222576050873 | 0.052370196524349044 |
| 0.4538768865847612 | 0.07085636337170394 | 0.053400683202913836 | 0.6427869803426212 | 0.052370196524349044 |
| 0.4297155255320146 | 0.07085636337170394 | 0.053400683202913836 | 0.6427869803426212 | 0.052370196524349044 |
| 0.42558388377505574 | 0.07085636337170394 | 0.053400683202913836 | 0.6328905221979035 | 0.052370196524349044 |
| 0.37909817365337545 | 0.07085636337170394 | 0.053400683202913836 | 0.6328905221979035 | 0.052370196524349044 |
| 0.3645473342101959 | 0.07085636337170394 | 0.053400683202913836 | 0.6328905221979035 | 0.052370196524349044 |
| 0.36262055396206444 | 0.07085636337170394 | 0.053400683202913836 | 0.6044532261016083 | 0.052370196524349044 |
| 0.3585798533491954 | 0.07085636337170394 | 0.04490255045469805 | 0.6044532261016083 | 0.047205873794830336 |
| 0.3538345288232321 | 0.06476818068785488 | 0.040204414262693715 | 0.6044532261016083 | 0.037394482643962355 |
| 0.34112341487223097 | 0.056976518189368115 | 0.040204414262693715 | 0.6044532261016083 | 0.03001955369125982 |
| 0.27764469545794945 | 0.05363070285608441 | 0.040204414262693715 | 0.6044532261016083 | 0.02885877403778891 |
| 0.27764469545794945 | 0.05237320079667769 | 0.040204414262693715 | 0.6044532261016083 | 0.02885877403778891 |
| 0.24466784630296584 | 0.04968901214595009 | 0.040204414262693715 | 0.6044532261016083 | 0.02885877403778891 |
| 0.2139348000837406 | 0.04968901214595009 | 0.040204414262693715 | 0.6044532261016083 | 0.02885877403778891 |
| 0.2139348000837406 | 0.04968901214595009 | 0.040204414262693715 | 0.6044532261016083 | 0.02885877403778891 |
| 0.2139348000837406 | 0.04968901214595009 | 0.04490255045469805 | 0.6389952771571914 | 0.02885877403778891 |
| 0.2139348000837406 | 0.04968901214595009 | 0.053400683202913836 | 0.6427869803426212 | 0.02885877403778891 |
| 0.2139348000837406 | 0.04968901214595009 | 0.053400683202913836 | 0.6458222576050873 | 0.02885877403778891 |
| 0.24466784630296584 | 0.07085636337170394 | 0.18355486471250076 | 0.7366544734706596 | 0.052370196524349044 |

**Supplementary Table 4:**
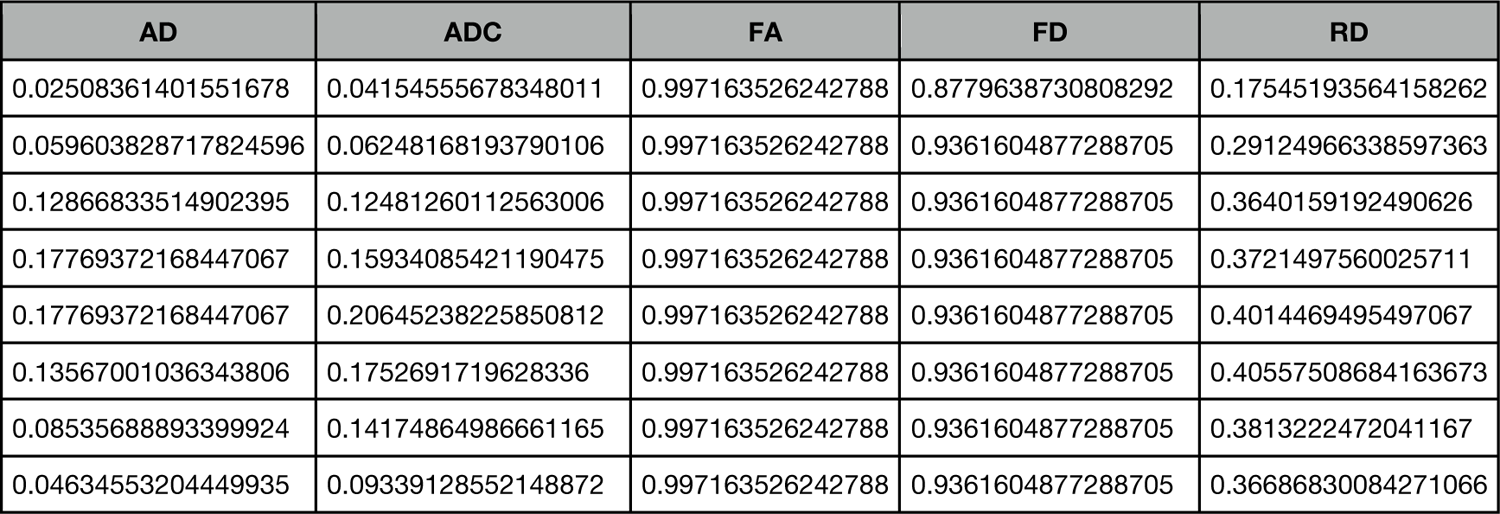
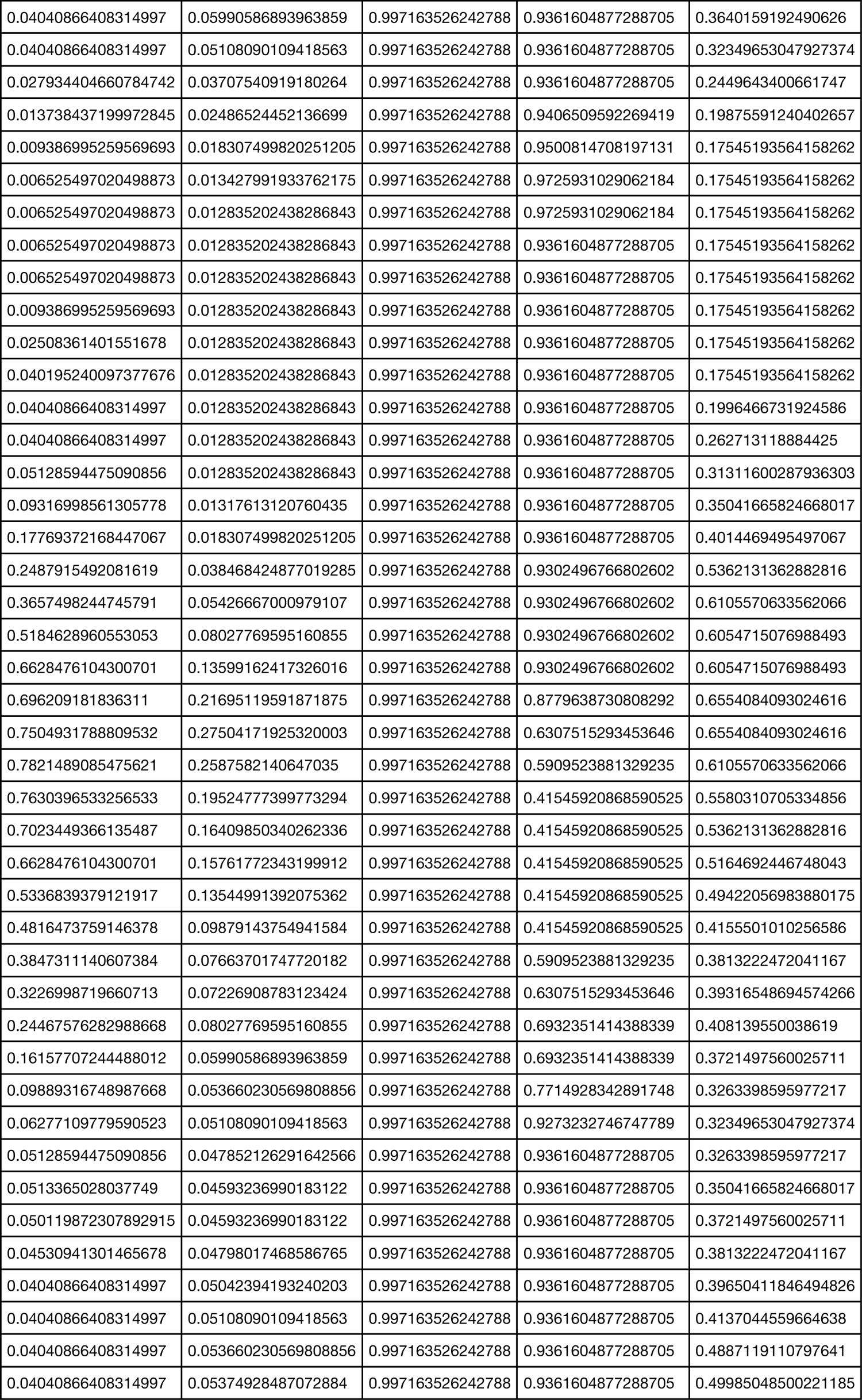
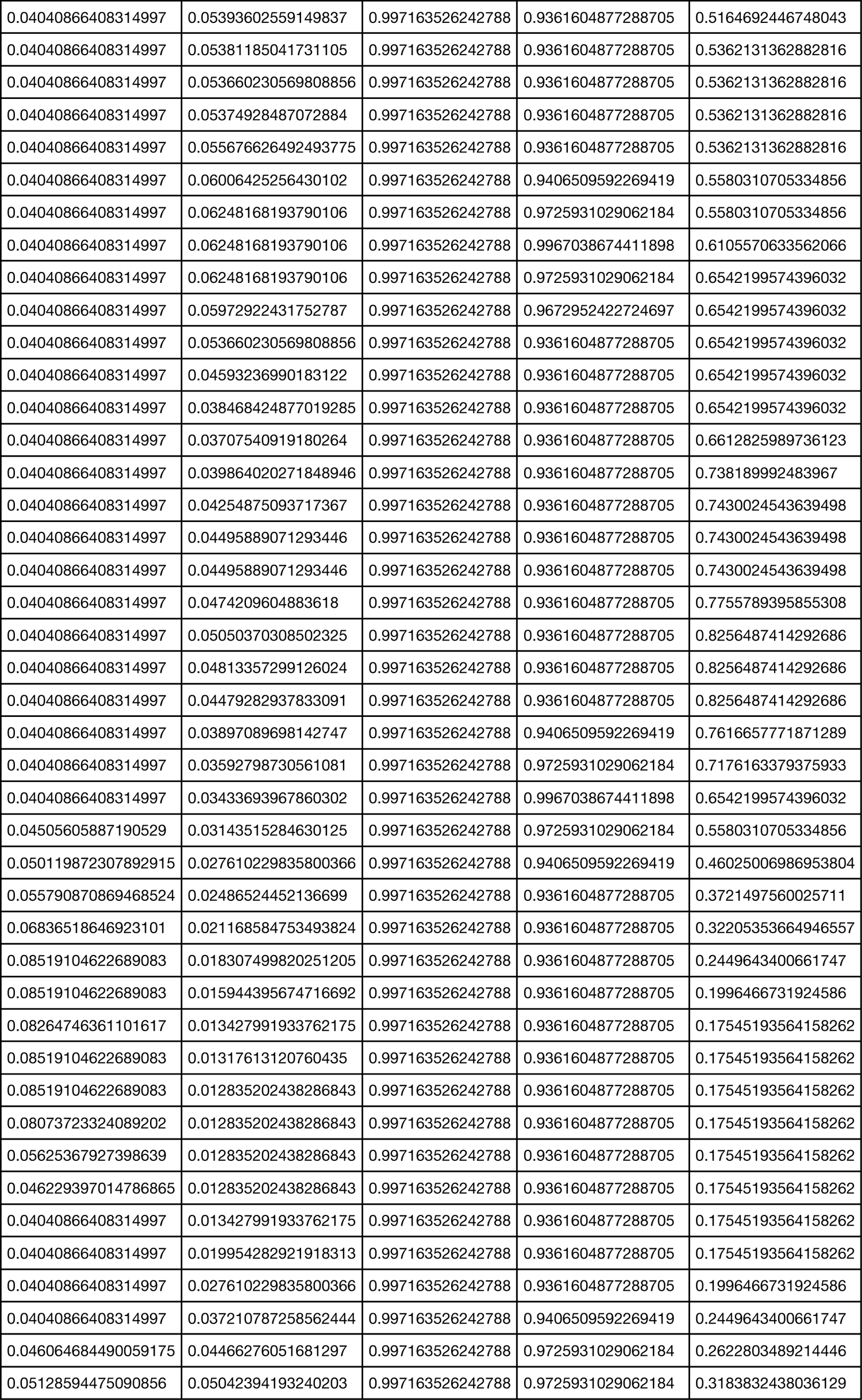
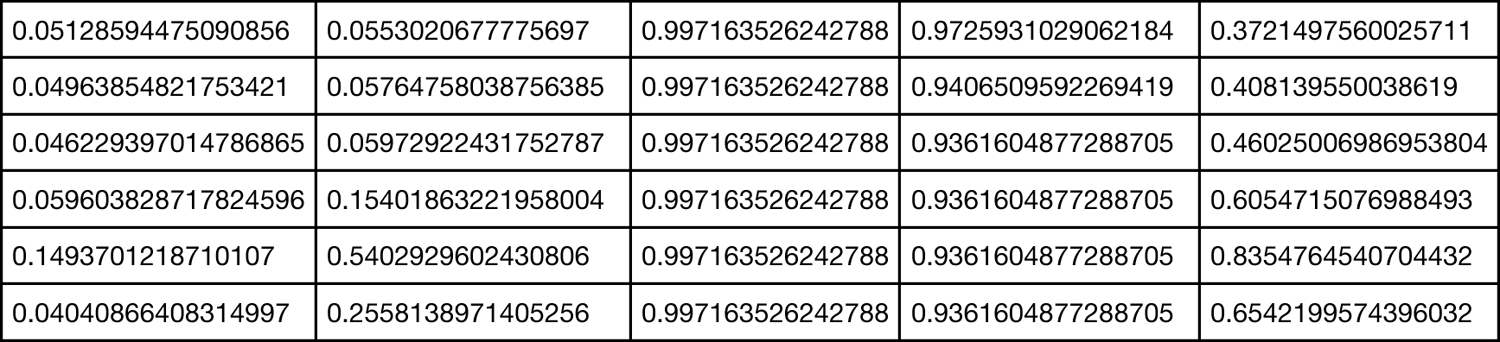
Corrected *p*-values (FDR-BH) along contralesional CST profiles

| AD | ADC | FA | FD | RD |
| --- | --- | --- | --- | --- |
| 0.02508361401551678 | 0.04154555678348011 | 0.997163526242788 | 0.8779638730808292 | 0.17545193564158262 |
| 0.059603828717824596 | 0.06248168193790106 | 0.997163526242788 | 0.9361604877288705 | 0.29124966338597363 |
| 0.12866833514902395 | 0.12481260112563006 | 0.997163526242788 | 0.9361604877288705 | 0.3640159192490626 |
| 0.17769372168447067 | 0.15934085421190475 | 0.997163526242788 | 0.9361604877288705 | 0.3721497560025711 |
| 0.17769372168447067 | 0.20645238225850812 | 0.997163526242788 | 0.9361604877288705 | 0.4014469495497067 |
| 0.13567001036343806 | 0.1752691719628336 | 0.997163526242788 | 0.9361604877288705 | 0.40557508684163673 |
| 0.08535688893399924 | 0.14174864986661165 | 0.997163526242788 | 0.9361604877288705 | 0.3813222472041167 |
| 0.04634553204449935 | 0.09339128552148872 | 0.997163526242788 | 0.9361604877288705 | 0.36686830084271066 |
| 0.04040866408314997 | 0.05990586893963859 | 0.997163526242788 | 0.9361604877288705 | 0.3640159192490626 |
| 0.04040866408314997 | 0.05108090109418563 | 0.997163526242788 | 0.9361604877288705 | 0.32349653047927374 |
| 0.027934404660784742 | 0.03707540919180264 | 0.997163526242788 | 0.9361604877288705 | 0.2449643400661747 |
| 0.013738437199972845 | 0.02486524452136699 | 0.997163526242788 | 0.9406509592269419 | 0.19875591240402657 |
| 0.009386995259569693 | 0.018307499820251205 | 0.997163526242788 | 0.9500814708197131 | 0.17545193564158262 |
| 0.006525497020498873 | 0.013427991933762175 | 0.997163526242788 | 0.9725931029062184 | 0.17545193564158262 |
| 0.006525497020498873 | 0.012835202438286843 | 0.997163526242788 | 0.9725931029062184 | 0.17545193564158262 |
| 0.006525497020498873 | 0.012835202438286843 | 0.997163526242788 | 0.9361604877288705 | 0.17545193564158262 |
| 0.006525497020498873 | 0.012835202438286843 | 0.997163526242788 | 0.9361604877288705 | 0.17545193564158262 |
| 0.009386995259569693 | 0.012835202438286843 | 0.997163526242788 | 0.9361604877288705 | 0.17545193564158262 |
| 0.02508361401551678 | 0.012835202438286843 | 0.997163526242788 | 0.9361604877288705 | 0.17545193564158262 |
| 0.040195240097377676 | 0.012835202438286843 | 0.997163526242788 | 0.9361604877288705 | 0.17545193564158262 |
| 0.04040866408314997 | 0.012835202438286843 | 0.997163526242788 | 0.9361604877288705 | 0.1996466731924586 |
| 0.04040866408314997 | 0.012835202438286843 | 0.997163526242788 | 0.9361604877288705 | 0.262713118884425 |
| 0.05128594475090856 | 0.012835202438286843 | 0.997163526242788 | 0.9361604877288705 | 0.31311600287936303 |
| 0.09316998561305778 | 0.01317613120760435 | 0.997163526242788 | 0.9361604877288705 | 0.35041665824668017 |
| 0.17769372168447067 | 0.018307499820251205 | 0.997163526242788 | 0.9361604877288705 | 0.4014469495497067 |
| 0.2487915492081619 | 0.038468424877019285 | 0.997163526242788 | 0.9302496766802602 | 0.5362131362882816 |
| 0.3657498244745791 | 0.05426667000979107 | 0.997163526242788 | 0.9302496766802602 | 0.6105570633562066 |
| 0.5184628960553053 | 0.08027769595160855 | 0.997163526242788 | 0.9302496766802602 | 0.6054715076988493 |
| 0.6628476104300701 | 0.13599162417326016 | 0.997163526242788 | 0.9302496766802602 | 0.6054715076988493 |
| 0.696209181836311 | 0.21695119591871875 | 0.997163526242788 | 0.8779638730808292 | 0.6554084093024616 |
| 0.7504931788809532 | 0.27504171925320003 | 0.997163526242788 | 0.6307515293453646 | 0.6554084093024616 |
| 0.7821489085475621 | 0.2587582140647035 | 0.997163526242788 | 0.5909523881329235 | 0.6105570633562066 |
| 0.7630396533256533 | 0.19524777399773294 | 0.997163526242788 | 0.41545920868590525 | 0.5580310705334856 |
| 0.7023449366135487 | 0.16409850340262336 | 0.997163526242788 | 0.41545920868590525 | 0.5362131362882816 |
| 0.6628476104300701 | 0.15761772343199912 | 0.997163526242788 | 0.41545920868590525 | 0.5164692446748043 |
| 0.5336839379121917 | 0.13544991392075362 | 0.997163526242788 | 0.41545920868590525 | 0.49422056983880175 |
| 0.4816473759146378 | 0.09879143754941584 | 0.997163526242788 | 0.41545920868590525 | 0.4155501010256586 |
| 0.3847311140607384 | 0.07663701747720182 | 0.997163526242788 | 0.5909523881329235 | 0.3813222472041167 |
| 0.3226998719660713 | 0.07226908783123424 | 0.997163526242788 | 0.6307515293453646 | 0.39316548694574266 |
| 0.24467576282988668 | 0.08027769595160855 | 0.997163526242788 | 0.6932351414388339 | 0.408139550038619 |
| 0.16157707244488012 | 0.05990586893963859 | 0.997163526242788 | 0.6932351414388339 | 0.3721497560025711 |
| 0.09889316748987668 | 0.053660230569808856 | 0.997163526242788 | 0.7714928342891748 | 0.3263398595977217 |
| 0.06277109779590523 | 0.05108090109418563 | 0.997163526242788 | 0.9273232746747789 | 0.32349653047927374 |
| 0.05128594475090856 | 0.047852126291642566 | 0.997163526242788 | 0.9361604877288705 | 0.3263398595977217 |
| 0.0513365028037749 | 0.04593236990183122 | 0.997163526242788 | 0.9361604877288705 | 0.35041665824668017 |
| 0.050119872307892915 | 0.04593236990183122 | 0.997163526242788 | 0.9361604877288705 | 0.3721497560025711 |
| 0.04530941301465678 | 0.04798017468586765 | 0.997163526242788 | 0.9361604877288705 | 0.3813222472041167 |
| 0.04040866408314997 | 0.05042394193240203 | 0.997163526242788 | 0.9361604877288705 | 0.39650411846494826 |
| 0.04040866408314997 | 0.05108090109418563 | 0.997163526242788 | 0.9361604877288705 | 0.4137044559664638 |
| 0.04040866408314997 | 0.053660230569808856 | 0.997163526242788 | 0.9361604877288705 | 0.4887119110797641 |
| 0.04040866408314997 | 0.05374928487072884 | 0.997163526242788 | 0.9361604877288705 | 0.49985048500221185 |
| 0.04040866408314997 | 0.05393602559149837 | 0.997163526242788 | 0.9361604877288705 | 0.5164692446748043 |
| 0.04040866408314997 | 0.05381185041731105 | 0.997163526242788 | 0.9361604877288705 | 0.5362131362882816 |
| 0.04040866408314997 | 0.053660230569808856 | 0.997163526242788 | 0.9361604877288705 | 0.5362131362882816 |
| 0.04040866408314997 | 0.05374928487072884 | 0.997163526242788 | 0.9361604877288705 | 0.5362131362882816 |
| 0.04040866408314997 | 0.055676626492493775 | 0.997163526242788 | 0.9361604877288705 | 0.5362131362882816 |
| 0.04040866408314997 | 0.06006425256430102 | 0.997163526242788 | 0.9406509592269419 | 0.5580310705334856 |
| 0.04040866408314997 | 0.06248168193790106 | 0.997163526242788 | 0.9725931029062184 | 0.5580310705334856 |
| 0.04040866408314997 | 0.06248168193790106 | 0.997163526242788 | 0.9967038674411898 | 0.6105570633562066 |
| 0.04040866408314997 | 0.06248168193790106 | 0.997163526242788 | 0.9725931029062184 | 0.6542199574396032 |
| 0.04040866408314997 | 0.05972922431752787 | 0.997163526242788 | 0.9672952422724697 | 0.6542199574396032 |
| 0.04040866408314997 | 0.053660230569808856 | 0.997163526242788 | 0.9361604877288705 | 0.6542199574396032 |
| 0.04040866408314997 | 0.04593236990183122 | 0.997163526242788 | 0.9361604877288705 | 0.6542199574396032 |
| 0.04040866408314997 | 0.038468424877019285 | 0.997163526242788 | 0.9361604877288705 | 0.6542199574396032 |
| 0.04040866408314997 | 0.03707540919180264 | 0.997163526242788 | 0.9361604877288705 | 0.6612825989736123 |
| 0.04040866408314997 | 0.039864020271848946 | 0.997163526242788 | 0.9361604877288705 | 0.738189992483967 |
| 0.04040866408314997 | 0.04254875093717367 | 0.997163526242788 | 0.9361604877288705 | 0.7430024543639498 |
| 0.04040866408314997 | 0.04495889071293446 | 0.997163526242788 | 0.9361604877288705 | 0.7430024543639498 |
| 0.04040866408314997 | 0.04495889071293446 | 0.997163526242788 | 0.9361604877288705 | 0.7430024543639498 |
| 0.04040866408314997 | 0.0474209604883618 | 0.997163526242788 | 0.9361604877288705 | 0.7755789395855308 |
| 0.04040866408314997 | 0.05050370308502325 | 0.997163526242788 | 0.9361604877288705 | 0.8256487414292686 |
| 0.04040866408314997 | 0.04813357299126024 | 0.997163526242788 | 0.9361604877288705 | 0.8256487414292686 |
| 0.04040866408314997 | 0.04479282937833091 | 0.997163526242788 | 0.9361604877288705 | 0.8256487414292686 |
| 0.04040866408314997 | 0.03897089698142747 | 0.997163526242788 | 0.9406509592269419 | 0.7616657771871289 |
| 0.04040866408314997 | 0.03592798730561081 | 0.997163526242788 | 0.9725931029062184 | 0.7176163379375933 |
| 0.04040866408314997 | 0.03433693967860302 | 0.997163526242788 | 0.9967038674411898 | 0.6542199574396032 |
| 0.04505605887190529 | 0.03143515284630125 | 0.997163526242788 | 0.9725931029062184 | 0.5580310705334856 |
| 0.050119872307892915 | 0.027610229835800366 | 0.997163526242788 | 0.9406509592269419 | 0.46025006986953804 |
| 0.055790870869468524 | 0.02486524452136699 | 0.997163526242788 | 0.9361604877288705 | 0.3721497560025711 |
| 0.06836518646923101 | 0.021168584753493824 | 0.997163526242788 | 0.9361604877288705 | 0.32205353664946557 |
| 0.08519104622689083 | 0.018307499820251205 | 0.997163526242788 | 0.9361604877288705 | 0.2449643400661747 |
| 0.08519104622689083 | 0.015944395674716692 | 0.997163526242788 | 0.9361604877288705 | 0.1996466731924586 |
| 0.08264746361101617 | 0.013427991933762175 | 0.997163526242788 | 0.9361604877288705 | 0.17545193564158262 |
| 0.08519104622689083 | 0.01317613120760435 | 0.997163526242788 | 0.9361604877288705 | 0.17545193564158262 |
| 0.08519104622689083 | 0.012835202438286843 | 0.997163526242788 | 0.9361604877288705 | 0.17545193564158262 |
| 0.08073723324089202 | 0.012835202438286843 | 0.997163526242788 | 0.9361604877288705 | 0.17545193564158262 |
| 0.05625367927398639 | 0.012835202438286843 | 0.997163526242788 | 0.9361604877288705 | 0.17545193564158262 |
| 0.046229397014786865 | 0.012835202438286843 | 0.997163526242788 | 0.9361604877288705 | 0.17545193564158262 |
| 0.04040866408314997 | 0.013427991933762175 | 0.997163526242788 | 0.9361604877288705 | 0.17545193564158262 |
| 0.04040866408314997 | 0.019954282921918313 | 0.997163526242788 | 0.9361604877288705 | 0.17545193564158262 |
| 0.04040866408314997 | 0.027610229835800366 | 0.997163526242788 | 0.9361604877288705 | 0.1996466731924586 |
| 0.04040866408314997 | 0.037210787258562444 | 0.997163526242788 | 0.9406509592269419 | 0.2449643400661747 |
| 0.046064684490059175 | 0.04466276051681297 | 0.997163526242788 | 0.9725931029062184 | 0.2622803489214446 |
| 0.05128594475090856 | 0.05042394193240203 | 0.997163526242788 | 0.9725931029062184 | 0.3183832438036129 |
| 0.05128594475090856 | 0.0553020677775697 | 0.997163526242788 | 0.9725931029062184 | 0.3721497560025711 |
| 0.04963854821753421 | 0.05764758038756385 | 0.997163526242788 | 0.9406509592269419 | 0.408139550038619 |
| 0.046229397014786865 | 0.05972922431752787 | 0.997163526242788 | 0.9361604877288705 | 0.46025006986953804 |
| 0.059603828717824596 | 0.15401863221958004 | 0.997163526242788 | 0.9361604877288705 | 0.6054715076988493 |
| 0.1493701218710107 | 0.5402929602430806 | 0.997163526242788 | 0.9361604877288705 | 0.8354764540704432 |
| 0.04040866408314997 | 0.2558138971405256 | 0.997163526242788 | 0.9361604877288705 | 0.6542199574396032 |

**Supplementary Table 5a:** *p*-values for histogram-based features of ipsilesional tract profiles (M, STD, KU and SK)

|  | Mean | Standard_deviation | Kurtosis | Skewness |
| --- | --- | --- | --- | --- |
| **AD** |  |  |  |  |
| **Pvalue** | 0.00035915874512317827 | 0.2341572786697913 | 0.366947543185421 | 0.36481618144418837 |
| **Statistic** | 1000.0 | 1469.0 | 1537.0 | 1536.0 |
| **ADC** |  |  |  |  |
| **Pvalue** | 0.00020793562410994644 | 0.0006843192729760747 | 0.06445201298720575 | 0.38847701594380935 |
| **Statistic** | 974.0 | 1032.0 | 1329.0 | 1547.0 |
| **FA** |  |  |  |  |
| **Pvalue** | 0.015003176742231243 | 0.347920379642365 | 0.0007117261438705842 | 0.1485877267971279 |
| **Statistic** | 1214.0 | 1528.0 | 1034.0 | 1413.0 |
| **FD** |  |  |  |  |
| **Pvalue** | 0.07190077569372236 | 0.2482846512862143 | 0.36481618144418837 | 0.08694737148719373 |
| **Statistic** | 1339.0 | 1477.0 | 1536.0 | 1357.0 |
| **RD** |  |  |  |  |
| **Pvalue** | 0.00038203487598156724 | 0.001288235588776543 | 0.10629430993736705 | 0.4125591732724999 |
| **Statistic** | 1003.0 | 1065.0 | 1377.0 | 1558.0 |

**Supplementary Table 5b:** p-values for histogram-based features of contralesional tract profiles (M, STD, KU and SK)

|  | Mean | Standard_deviation | Kurtosis | Skewness |
| --- | --- | --- | --- | --- |
| **AD** |  |  |  |  |
| **Pvalue** | 0.0029473206024855916 | 0.10944566682997958 | 0.3190837409993945 | 0.11375107562409548 |
| **Statistic** | 1111.0 | 1380.0 | 1514.0 | 1384.0 |
| **ADC** |  |  |  |  |
| **Pvalue** | 0.001288235588776543 | 0.0816905028870411 | 0.14085083793248715 | 0.09151784128287349 |
| **Statistic** | 1065.0 | 1351.0 | 1407.0 | 1362.0 |
| **FA** |  |  |  |  |
| **Pvalue** | 0.4302883040679208 | 0.3030707431663115 | 0.10525856321503085 | 0.3375093416727035 |
| **Statistic** | 1566.0 | 1506.0 | 1376.0 | 1523.0 |
| **FD** |  |  |  |  |
| **Pvalue** | 0.47966582022590476 | 0.30505263433040164 | 0.18534049210053155 | 0.02632941924235198 |
| **Statistic** | 1588.0 | 1507.0 | 1439.0 | 1255.0 |
| **RD** |  |  |  |  |
| **Pvalue** | 0.016339233313890682 | 0.05018258478215847 | 0.14727888645121695 | 0.12857297507759896 |
| **Statistic** | 1220.0 | 1307.0 | 1412.0 | 1397.0 |

## Notes

### Competing Interest Statement

The authors have declared no competing interest.

### Author Declarations

The study proposal is in accordance with ethical standards of the Declaration of Helsinki and was approved by the Ethics Commission of the Charite University Hospital (#EA1/016/19). All patients provided written informed consent for medical evaluations and treatments within the scope of the study.

## References

1. Weller, M. et al. Glioma. Nat. Rev. Dis. Prim. 1, 15017 (2015).

2. Basser, P. J., Mattiello, J. & LeBihan, D. MR diffusion tensor spectroscopy and imaging. Biophys. J. 66, 259–267 (1994).

3. Le Bihan, D. & Johansen-Berg, H. Diffusion MRI at 25: exploring brain tissue structure and function. Neuroimage 61, 324–341 (2012).

4. Bartsch, A. J., Biller, A. & Homola, G. A. Chapter 23 - Presurgical Tractography Applications. in (eds. Johansen-Berg, H. & Behrens, T. E. J. B. T.-D. M. R. I. (Second E.) 531–567 (Academic Press, 2014). doi: https://doi.org/10.1016/B978-0-12-396460-1.00023-8.

5. Mori, S. Chapter 9 - Three-dimensional tract reconstruction. in (ed. Mori, S. B. T.-I. to D. T. I.) 93–123 (Elsevier Science B.V., 2007). doi:https://doi.org/10.1016/B978-044452828-5/50023-5.

6. Pujol, S. Chapter 4 - Imaging White Matter Anatomy for Brain Tumor Surgery. in (ed. Golby, A. J. B. T.-I.-G. N.) 91–121 (Academic Press, 2015). doi:https://doi.org/10.1016/B978-0-12-800870-6.00004-2.

7. Dell’Acqua, F. & Tournier, J.-D. Modelling white matter with spherical deconvolution: How and why? NMR Biomed. 32, e3945 (2019).

8. Jeurissen, B., Descoteaux, M., Mori, S. & Leemans, A. Diffusion MRI fiber tractography of the brain. NMR Biomed. 32, e3785 (2019).

9. Essayed, W. I. et al. White matter tractography for neurosurgical planning: A topography-based review of the current state of the art. NeuroImage Clin. 15, 659– 672 (2017).

10. Romano, A. et al. Pre-surgical planning and MR-tractography utility in brain tumour resection. Eur. Radiol. 19, 2798–2808 (2009).

11. D’Souza, S., Ormond, D. R., Costabile, J. & Thompson, J. A. Fiber-tract localized diffusion coefficients highlight patterns of white matter disruption induced by proximity to glioma. PLoS One 14, e0225323 (2019).

12. Soares, J., Marques, P., Alves, V. & Sousa, N. A hitchhiker’s guide to diffusion tensor imaging. Frontiers in Neuroscience vol. 7 31 (2013).

13. Rosenstock, T. et al. Specific DTI seeding and diffusivity-analysis improve the quality and prognostic value of TMS-based deterministic DTI of the pyramidal tract. NeuroImage Clin. 16, 276–285 (2017).

14. Fekonja, L. S. et al. Detecting Corticospinal Tract Impairment in Tumor Patients With Fiber Density and Tensor-Based Metrics. Frontiers in Oncology vol. 10 3256 (2021).

15. Bells, S. et al. Tractometry-comprehensive multi-modal quantitative assessment of white matter along specific tracts. Proc. ISMRM 2011 (2011).

16. Colby, J. B. et al. Along-tract statistics allow for enhanced tractography analysis. Neuroimage 59, 3227–3242 (2012).

17. Yeatman, J. D., Dougherty, R. F., Myall, N. J., Wandell, B. A. & Feldman, H. M. Tract Profiles of White Matter Properties: Automating Fiber-Tract Quantification. PLoS One 7, e49790 (2012).

18. O’Donnell, L. J., Westin, C.-F. & Golby, A. J. Tract-based morphometry for white matter group analysis. Neuroimage 45, 832–844 (2009).

19. Yendiki, A. et al. Automated Probabilistic Reconstruction of White-Matter Pathways in Health and Disease Using an Atlas of the Underlying Anatomy. Frontiers in Neuroinformatics vol. 5 23 (2011).

20. Jeurissen, B., Leemans, A., Tournier, J.-D., Jones, D. K. & Sijbers, J. Investigating the prevalence of complex fiber configurations in white matter tissue with diffusion magnetic resonance imaging. Hum. Brain Mapp. 34, 2747–2766 (2013).

21. Riffert, T. W., Schreiber, J., Anwander, A. & Knösche, T. R. Beyond fractional anisotropy: extraction of bundle-specific structural metrics from crossing fiber models. Neuroimage 100, 176–191 (2014).

22. Vos, S. B., Jones, D. K., Jeurissen, B., Viergever, M. A. & Leemans, A. The influence of complex white matter architecture on the mean diffusivity in diffusion tensor MRI of the human brain. Neuroimage 59, 2208–2216 (2012).

23. Tournier, J.-D. Diffusion MRI in the brain–Theory and concepts. Prog. Nucl. Magn. Reson. Spectrosc. 112, 1–16 (2019).

24. Roine, T. et al. Isotropic non-white matter partial volume effects in constrained spherical deconvolution. Front. Neuroinform. 8, 28 (2014).

25. Dhollander, T., Raffelt, D. & Connelly, A. Unsupervised 3-tissue response function estimation from single-shell or multi-shell diffusion MR data without a co-registered T1 image. in ISMRM Workshop on Breaking the Barriers of Diffusion MRI vol. 5 5 (2016).

26. Roine, T. et al. Informed constrained spherical deconvolution (iCSD). Med. Image Anal. 24, 269–281 (2015).

27. Rasmussen, P. M., Madsen, K. H., Lund, T. E. & Hansen, L. K. Visualization of nonlinear kernel models in neuroimaging by sensitivity maps. Neuroimage 55, 1120–1131 (2011).

28. Jeurissen, B., Tournier, J.-D., Dhollander, T., Connelly, A. & Sijbers, J. Multi-tissue constrained spherical deconvolution for improved analysis of multi-shell diffusion MRI data. Neuroimage 103, 411–426 (2014).

29. Tournier, J.-D., Calamante, F., Gadian, D. G. & Connelly, A. Direct estimation of the fiber orientation density function from diffusion-weighted MRI data using spherical deconvolution. Neuroimage 23, 1176–1185 (2004).

30. Tournier, J.-D., Mori, S. & Leemans, A. Diffusion tensor imaging and beyond. Magn. Reson. Med. 65, 1532–1556 (2011).

31. Mormina, E. et al. MRI tractography of corticospinal tract and arcuate fasciculus in high-grade gliomas performed by constrained spherical deconvolution: qualitative and quantitative analysis. *Am*. J. Neuroradiol. 36, 1853–1858 (2015).

32. Raffelt, D. A. et al. Investigating white matter fibre density and morphology using fixel-based analysis. Neuroimage 144, 58–73 (2017).

33. Dhollander, T. et al. Fixel-based Analysis of Diffusion MRI: Methods, Applications, Challenges and Opportunities. Neuroimage 118417 (2021) doi: https://doi.org/10.1016/j.neuroimage.2021.118417.

34. Cortes, C. & Vapnik, V. Support-vector networks. Mach. Learn. 20, 273–297 (1995).

35. Noble, W. S. What is a support vector machine? Nat. Biotechnol. 24, 1565–1567 (2006).

36. Bzdok, D., Krzywinski, M. & Altman, N. Machine learning: supervised methods. Nat. Methods 15, 5–6 (2018).

37. Chen, X. & Jeong, J. C. Enhanced recursive feature elimination. in Sixth International Conference on Machine Learning and Applications (ICMLA 2007) 429–435 (2007). doi:10.1109/ICMLA.2007.35.

38. Jain, A. & Zongker, D. Feature selection: evaluation, application, and small sample performance. IEEE Trans. Pattern Anal. Mach. Intell. 19, 153–158 (1997).

39. Jolliffe, I. Principal Component Analysis BT - International Encyclopedia of Statistical Science. in (ed. Lovric, M.) 1094–1096 (Springer Berlin Heidelberg, 2011). doi:10.1007/978-3-642-04898-2_455.

40. Hotelling, H. Analysis of a complex of statistical variables into principal components. J. Educ. Psychol. 24, 417–441 (1933).

41. Alexander, A. L., Lee, J. E., Lazar, M. & Field, A. S. Diffusion tensor imaging of the brain. Neurotherapeutics 4, 316–329 (2007).

42. Harsan, L. A. et al. Brain dysmyelination and recovery assessment by noninvasive in vivo diffusion tensor magnetic resonance imaging. J. Neurosci. Res. 83, 392–402 (2006).

43. Sun, S.-W. et al. Noninvasive detection of cuprizone induced axonal damage and demyelination in the mouse corpus callosum. Magn. Reson. Med. 55, 302–308 (2006).

44. Concha, L. A macroscopic view of microstructure: Using diffusion-weighted images to infer damage, repair, and plasticity of white matter. Neuroscience 276, 14–28 (2014).

45. Liu, D. et al. Alterations of white matter integrity associated with cognitive deficits in patients with glioma. Brain Behav. 10, e01639 (2020).

46. Lo, A., Chernoff, H., Zheng, T. & Lo, S.-H. Why significant variables aren’t automatically good predictors. Proc. Natl. Acad. Sci. U. S. A. 112, 13892–13897 (2015).

47. Bzdok, D., Engemann, D. & Thirion, B. Inference and Prediction Diverge in Biomedicine. Patterns 1, (2020).

48. Richie-Halford, A., Yeatman, J. D., Simon, N. & Rokem, A. Multidimensional analysis and detection of informative features in human brain white matter. PLOS Comput. Biol. 17, e1009136 (2021).

49. Huber, E., Henriques, R. N., Owen, J. P., Rokem, A. & Yeatman, J. D. Applying microstructural models to understand the role of white matter in cognitive development. Dev. Cogn. Neurosci. 36, 100624 (2019).

50. Tian, L. & Ma, L. Microstructural Changes of the Human Brain from Early to Mid-Adulthood. Frontiers in Human Neuroscience vol. 11 393 (2017).

51. Lövdén, M. et al. Changes in perceptual speed and white matter microstructure in the corticospinal tract are associated in very old age. Neuroimage 102, 520–530 (2014).

52. Salat, D. H. et al. Age-related alterations in white matter microstructure measured by diffusion tensor imaging. Neurobiol. Aging 26, 1215–1227 (2005).

53. Schilling, K. G. et al. Limits to anatomical accuracy of diffusion tractography using modern approaches. Neuroimage 185, 1–11 (2019).

54. Maier-Hein, K. H. et al. The challenge of mapping the human connectome based on diffusion tractography. Nat. Commun. 8, 1349 (2017).

55. Aydogan, D. B. et al. When tractography meets tracer injections: a systematic study of trends and variation sources of diffusion-based connectivity. Brain Struct. Funct. 223, 2841–2858 (2018).

56. Tournier, J.-D. et al. MRtrix3: A fast, flexible and open software framework for medical image processing and visualisation. Neuroimage 202, 116137 (2019).

57. Raffelt, D. et al. Apparent Fibre Density: A novel measure for the analysis of diffusion-weighted magnetic resonance images. Neuroimage 59, 3976–3994 (2012).

58. Picht, T., Frey, D., Thieme, S., Kliesch, S. & Vajkoczy, P. Presurgical navigated TMS motor cortex mapping improves outcome in glioblastoma surgery: a controlled observational study. J. Neurooncol. 126, 535–543 (2016).

59. Avants, B. B. et al. A reproducible evaluation of ANTs similarity metric performance in brain image registration. Neuroimage 54, 2033–2044 (2011).

60. Avants, B. B. et al. The Insight ToolKit image registration framework. Frontiers in Neuroinformatics vol. 8 44 (2014).

61. Grabner, Gř. et al. Symmetric atlasing and model based segmentation: an application to the hippocampus in older adults. in International Conference on Medical Image Computing and Computer-Assisted Intervention 58–66 (Springer, 2006).

62. Veraart, J. et al. Denoising of diffusion MRI using random matrix theory. Neuroimage 142, 394–406 (2016).

63. Kellner, E., Dhital, B., Kiselev, V. G. & Reisert, M. Gibbs-ringing artifact removal based on local subvoxel-shifts. Magn. Reson. Med. 76, 1574–1581 (2016).

64. Leemans, A. & Jones, D. K. The B-matrix must be rotated when correcting for subject motion in DTI data. Magn. Reson. Med. An Off. J. Int. Soc. Magn. Reson. Med. 61, 1336–1349 (2009).

65. Andersson, J. L. R. et al. Towards a comprehensive framework for movement and distortion correction of diffusion MR images: Within volume movement. Neuroimage 152, 450–466 (2017).

66. Andersson, J. L. R., Skare, S. & Ashburner, J. How to correct susceptibility distortions in spin-echo echo-planar images: application to diffusion tensor imaging. Neuroimage 20, 870–888 (2003).

67. Jenkinson, M., Beckmann, C. F., Behrens, T. E. J., Woolrich, M. W. & Smith, S. M. Fsl. Neuroimage 62, 782–790 (2012).

68. Tustison, N. J. et al. N4ITK: improved N3 bias correction. IEEE Trans. Med. Imaging 29, 1310–1320 (2010).

69. Dyrby, T. B. et al. Interpolation of diffusion weighted imaging datasets. Neuroimage 103, 202–213 (2014).

70. Veraart, J., Sijbers, J., Sunaert, S., Leemans, A. & Jeurissen, B. Weighted linear least squares estimation of diffusion MRI parameters: strengths, limitations, and pitfalls. Neuroimage 81, 335–346 (2013).

71. Tournier, J.-D., Calamante, F. & Connelly, A. Robust determination of the fibre orientation distribution in diffusion MRI: non-negativity constrained super-resolved spherical deconvolution. Neuroimage 35, 1459–1472 (2007).

72. Tournier, J.-D., Calamante, F. & Connelly, A. Improved probabilistic streamlines tractography by 2 nd order integration over fibre orientation distributions. in (2009).

73. Mayka, M. A., Corcos, D. M., Leurgans, S. E. & Vaillancourt, D. E. Three-dimensional locations and boundaries of motor and premotor cortices as defined by functional brain imaging: A meta-analysis. Neuroimage 31, 1453–1474 (2006).

74. Virtanen, P. et al. SciPy 1.0: fundamental algorithms for scientific computing in Python. Nat. Methods 17, 261–272 (2020).

75. Waskom, M. et al. mwaskom/seaborn: v0.11.1 (December 2020). (2020) doi:10.5281/ZENODO.4379347.

76. Seabold, S. & Perktold, J. Statsmodels: Econometric and Statistical Modeling with Python. Proc. 9th Python Sci. Conf. 2010, (2010).

77. Hunter, J. D. Matplotlib: A 2D Graphics Environment. Comput. Sci. Eng. 9, 90–95 (2007).

78. Haynes, W. Benjamini–Hochberg Method BT - Encyclopedia of Systems Biology. in (eds. Dubitzky, W., Wolkenhauer, O., Cho, K.-H. & Yokota, H.) 78 (Springer New York, 2013). doi:10.1007/978-1-4419-9863-7_1215.

79. Hardoon, D. R., Szedmak, S. & Shawe-Taylor, J. Canonical correlation analysis: An overview with application to learning methods. Neural Comput. 16, 2639–2664 (2004).

80. Wang, Z. et al. Support vector machine based aphasia classification of transcranial magnetic stimulation language mapping in brain tumor patients. NeuroImage Clin. 29, 102536 (2021).

81. Gaonkar, B., T. Shinohara, R. & Davatzikos, C. Interpreting support vector machine models for multivariate group wise analysis in neuroimaging. Med. Image Anal. 24, 190–204 (2015).

82. Schölkopf, B., Smola, A. J. & Bach, F. Learning with kernels: support vector machines, regularization, optimization, and beyond. (MIT press, 2002).

83. Shawe-Taylor, J. & Cristianini, N. Kernel methods for pattern analysis. (Cambridge university press, 2004).

84. Murdoch, W. J., Singh, C., Kumbier, K., Abbasi-Asl, R. & Yu, B. Definitions, methods, and applications in interpretable machine learning. Proc. Natl. Acad. Sci. 116, 22071 LP – 22080 (2019).

85. Stewart, T. G., Zeng, D. & Wu, M. C. Constructing support vector machines with missing data. *WIREs Comput*. Stat. 10, e1430 (2018).

86. Batista, G. E. & Monard, M. C. A study of K-nearest neighbour as an imputation method. His 87, 48 (2002).

87. Lever, J., Krzywinski, M. & Altman, N. Points of significance: model selection and overfitting. (2016).

88. Petersen, M. L., Molinaro, A. M., Sinisi, S. E. & van der Laan, M. J. Cross-Validated Bagged Learning. J. Multivar. Anal. 25, 260–266 (2008).

